# Effect of regional deprivation on mental and physical health: a longitudinal natural experiment among refugees in Germany

**DOI:** 10.1101/2023.08.09.23293755

**Authors:** Louise Biddle, Kayvan Bozorgmehr

## Abstract

**Background:** Existing studies on contextual health effects struggle to account for selection bias, limiting causal interpretation. We use refugee dispersal in Germany as natural experiment to study the effect of small-area deprivation on mental and physical health, while considering the potential mediating role of housing and social context.

**Methods:** Refugees subject to dispersal (n=1400) are selected from a nation-wide longitudinal refugee study (IAB-SOEP-BAMF Panel; 2016-2018). Multi-level linear regression models, adjusted for age, sex, education, region of origin, federal state, asylum status and length of residence in Germany, are fitted to the change in mental and physical health subscales of the SF-12 depending on quintiles (Q1 – Q5) of district-level socioeconomic deprivation (German Index of Socio-Economic Deprivation, GISD). This is followed by mediation analyses (for housing and social context) and sensitivity analyses.

**Findings:** Residency in districts with moderate-high deprivation (Q4) has a negative impact on physical health (coef·: -2·2, 95%CI: -4·1;-0·2) compared to lowest deprivation (Q1). Moderate-high deprivation (Q4) also has a positive impact on mental health, but the effect is statistically insignificant following covariate adjustment (coef·: 2·5, 95%CI: -0·7;5·6). Comparisons with other deprivation quintiles are statistically insignificant.

**Interpretation:** The results point to gaps in health and social service provision for refugees living in the most deprived regions. Further efforts should be made to support integration of refugees into health and social systems in resource-poor regions, including improved interpreting services, specifically trained social workers and diversity-sensitive information offerings. Further research using longer timeframes and larger sample sizes are required to confirm results.

**Funding:** German Science Foundation (FOR: 2928/ GZ: BO5233/1-1).

## Panel1: Research in context

### Evidence before this study

The authors searched for keywords “context/deprivation/neighbourhood” and “health” in titles of systematic reviews indexed in PubMed to identify existing literature on the links between contextual characteristics and health, identifying 12 studies. Seminal studies have shown the impact of neighbourhood contextual characteristics on mortality, but also physical health outcomes such as self-rated health, cardiovascular disease, respiratory illness and health behaviours. Since the 1990s, the development of more sophisticated statistical methods in the field of social epidemiology allowed for the analysis of regional-level factors in individual-level health outcomes through the use of multi-level regression models. Such analyses have produced a more nuanced picture of the effects of deprivation on health, with some varied and/or inconclusive results for some health outcomes such as mental health. Despite the methodological benefits conferred by these approaches, issues with their interpretation remain: the decision to live in or move to a particular neighbourhood is invariably shaped by social, economic and cultural factors, which result in systematic differences between individuals in different regions (compositional bias). Natural experiments are a useful tool in disentangling compositional from contextual effects.

A recent systematic review examined studies analysing contextual effects on health using natural experiments among migrants. Existing studies from Denmark and Sweden use refugee dispersal processes as natural experiments, confirming the negative impacts of deprivation on physical health found in previous research, but showing mixed results for mental health. However, as these studies use register-based approaches with identification of refugees by nationality, they are subject to misclassification bias. Furthermore, they are carried out in contexts without residence requirements, and thus cannot ensure treatment adherence as secondary migration to other places of residence is possible after initial assignment.

### Added value of this study

This study uses refugee dispersal in Germany as natural experiment to study the causal effect of small-area deprivation on mental and physical health. The comprehensive refugee dispersal process in Germany (quasi-random dispersal at federal, regional and communal levels) and its associated 3-year residence rule provides an ideal policy environment to study contextual characteristics. The study uses survey data from a nation-wide longitudinal refugee study (IAB-SOEP-BAMF Panel; 2016-2018), meaning that the potential mediating role of social context and housing characteristics can be examined. It applies a difference-in-difference analysis, fitting multi-level linear regression models, adjusted for socio-demographic characteristics, to the change in mental and physical health subscales of the SF-12 depending on quintiles (Q1 – Q5) of district-level socioeconomic deprivation (German Index of Socio-Economic Deprivation, GISD). Residency in districts with moderate-high deprivation (Q4) has a negative impact on physical health (coef·: -2·2, 95%CI: -4·1;-0·2) when compared to living in districts with lowest deprivation (Q1). This result is robust to sensitivity analyses and not mediated by accommodation or social context variables. Residency in districts with moderate-high deprivation (Q4) has a positive impact on mental health, but this effect is not statistically significant in the fully adjusted model (coef·: 2·5, 95%CI: -0·7;5·6).

### Implications of all available evidence

The results of this study suggest that, in addition to affecting the development of ill health in the long term, small-area deprivation may exacerbate existing physical health issues by providing insufficient access to health and social services in the short term. Furthermore, the results of our study further contribute to the growing body of literature which shows complex effects of deprivation on mental health. While some studies have reported a negative impact of deprivation on symptoms of depression and anxiety, our study and other natural experiments using strict dispersal policies among refugees suggest that this may be due to selection effects. Robust evidence from other studies does, however, provide support for the negative effects of deprivation on psychiatric diagnoses and prescriptions.

## MANUSCRIPT

### Background

Understanding the health impacts of the places where people live has fascinated researchers for decades. Contextual factors of the place of residence can include such varied factors as the regional inequality, education, infrastructure development, green space, social capital or walkability ^1^. In social epidemiology, contextual factors are frequently operationalised as deprivation indices at small-area level. This allows for the joint assessments of multiple relevant factors which are often colinear. The effects of deprivation on health have been widely documented ^1–3^, but often suffer from compositional bias (Panel 1).

To overcome these issues, natural experiments are needed ^1^. Since the place of residence is not readily amenable to experimentation, situations where individuals are (quasi-)randomly distributed into neighbourhoods provide an opportunity to study contextual effects on health ^4^. The dispersal of refugees provides such opportunities, as it is organised in a quota-based system in several countries, allocating individuals to contexts at national or sub-national level based on factors such as population size or tax revenue, but independent of socio-demographic characteristics of the refugee population ^5^.

Refugee health is not a singularity and other marginalised populations may be subject to the same contextual exposures. Studying their health in natural experiments may thus serve as a lens, allowing us to explore the effects of deprivation for the health of other population groups. This must, however, be done with careful consideration of the causal mechanisms and potential mediators at play. In particular, previous studies have shown that the type of accommodation and accommodation size can have a direct impact on mental and physical health of refugees ^6,7^. The social context into which refugees are dispersed may further mediate the relationship between deprivation and health. Regions with lower deprivation may have more resources to invest in infrastructure conducive to social participation and engagement such as local parks, libraries, community centres and activity groups ^8^. Alternatively, areas of high deprivation may in fact be beneficial for migrant health through the existence of co-ethnic social networks which act as buffers for “acculturative stress” ^9,10^.

Given the above, the primary aim of this analysis is to investigate what impact living in an area of high deprivation has on the mental and physical health of refugees. The secondary aim is to assess whether social context and accommodation characteristics mediate the relationship between deprivation and health.

## Methods

This study employs a natural experiment design using longitudinal data from three waves of the IAB-SOEP-BAMF Refugee Panel (M3-M5; 2016-2018) in Germany^11^ to conduct a difference-in-difference analysis. Assignment to exposure (regional deprivation) is exogenous due to the allocation of refugees to different geographical contexts based on quasi-random administrative quotas.

### Study setting

Germany continues to host the highest number of refugees in Europe, with an estimated 2.1 million refugees residing in Germany in 2022 ^12^. Upon arrival in Germany, refugees are dispersed into communes based on a three-level dispersal process at federal, regional and communal levels (Supplementary File S1). While the dispersal of refugees is not entirely random, it remains exogenous as self-selection into communities by the refugees themselves is not possible. A further feature of the asylum system in Germany which makes it a unique natural experiment is the residence requirement policy (“Wohnsitzauflage”). The policy, enacted in several federal states, requires asylum seekers to remain resident in the commune to which they were assigned for the duration of their asylum application and up to 3 years following a decision.

### Data source

The IAB-SOEP-BAMF Panel ^11^ is an extension of the German socio-economic panel specifically tailored to the refugee population. The survey collects detailed information on social, economic, psychological and health indicators from a representative sample of refugees living in Germany. Sampling of participants is based on all refugees listed in the central register of foreign nationals (“Ausländerzentralregister”) who arrived in Germany between January 2013 and December 2016. The total adult sample (N=4855) consists of three waves (M3-M5), which were recruited between 2016 and 2017. The overall response rate was high at 48·7% ^13^. All waves were followed up in 2018 and thus have slightly different follow-up periods (1 vs. 2 years).

### Sample selection

In order to comply with criteria for the residence requirement policy, individuals were excluded who fulfilled one or more of the exemption criteria detailed in Supplementary File S2. The sample for this analysis hence includes a total of 1400 individuals, who were subject to the residence requirement policy at both t0 and t1 (n=1329), and those who are no longer subject to the policy at t1 (but were at t0) if they have not since moved to a different commune (n=71).

Analyses of the complete dataset show a loss follow-up of 51·6% and comparable rates across groups subject to the residence requirement policy and those who are not.

### Variables

Our primary outcome measures are change (t1-t0) in mental health score (mcs) and physical health score (pcs) derived from the SF-12. The scores are calculated using explorative factor analysis (PCA, varimax rotation) using the mean value of the SOEP 2004 population ^14^. Our exposure is regional deprivation in quintiles, with Q1 indicating lowest and Q5 highest deprivation. We use the 2012 German Index of Socioeconomic Deprivation (GISD) ^15^ on the level of communes. The GISD combines eight indicators on unemployment, education, income, tax revenue and debtors from the INKAR (indicators and maps on spatial and urban development in Germany and Europe) database using factor analysis ^15^.

Covariates were selected based on the directed acyclic graph (DAG) displayed in figure 1. Despite the natural experiment design, the uneven dispersal based on nationality at the national (and partly sub-national) level results in potential confounding through socio-demographic and asylum-related characteristics which need to be taken into account. In baseline models, we adjust for characteristics which influence the first-level dispersal process: federal state and region of origin. We use region of origin instead of nationality to avoid problems with empty cells. Region of origin groupings are based on the UN Geoscheme ^16^. Adjusting for region of origin also allows for potential differences in pre-migration experiences to be taken into account. Absolute mcs/pcs values at baseline assessment (t0) are also included in the baseline models. Given existing evidence on the uneven socio-demographic distribution of refugees across Germany ^17^, we further adjust for age, gender, highest educational attainment (as a proxy for individuals resources to navigate through the health and social system and health literacy aspects), asylum status and time since arrival in Germany.

**Figure 1:**
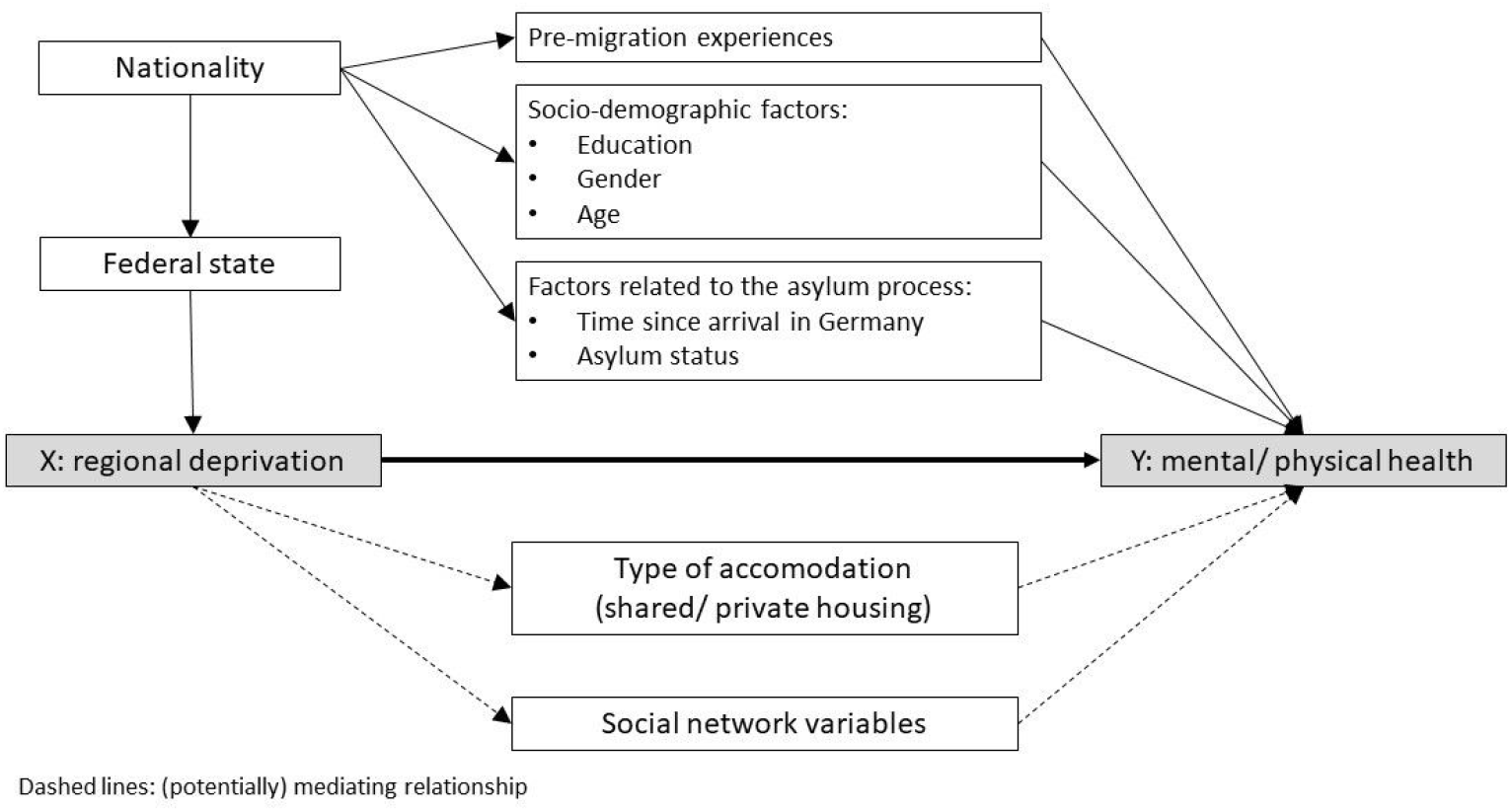
Directed Acyclic Graph depicting causal relationships between exposure, outcome and covariates used to guide the analysis.

The type of accommodation and the social context of participants are included in separate mediation analyses. Type of accommodation (private vs. shared) is included as a standalone variable. For the social context, variables on the size of the social network (number of new acquaintances since arriving in Germany from 1) Germany, 2) own country of origin and 3) other countries), the quality of the social network (feeling like an outsider, feeling socially isolated, feeling welcome, worries about xenophobia) and the perceived safety of the neighbourhood were included.

### Statistical methods

We use linear regression to model the relationship between regional deprivation in quintiles and change in mcs/pcs. Due to comparatively small sample sizes in the more deprived quintiles, robust variances were estimated using bootstrapping with 1000 replications on all models. Baseline and socio-demographic variables are introduced one at a time. For linear models, we check model fit using adjusted r2 as well as Wald tests on nested models to assess the relevance of newly included variables. Multicollinearity is assessed using variance inflation factors (VIF). Missing variables were handled by listwise deletion.

We then use multi-level models, fitting random intercepts at the level of the communes to account for clustering at the contextual level. Intra-class correlation (ICC) and likelihood ratio tests (clustered vs. linear models) are used to judge relevance of clustering; Akaike’s and Bayesian information criterion (AIC/BIC) are used to judge model fit. Proportional change in variance (PCV) is used to assess the relevance of individual-vs. contextual-level variables in explaining the variation in outcome between clusters. Further mediation and sensitivity analyses are carried out with multi-level models if there is substantial evidence for clustering (LR-test of clustered vs. linear model p<0·05) or on linear models if there is no evidence of clustering in order to avoid unnecessarily inflating estimate precision (these are deemed to be the “final models”).

For the analysis of mediation through social network variables, relevant variables are first chosen by independently introducing variables into the final model and assessing the relevance of variables based on the Wald test (p<0·05). All variables deemed relevant were then included in a “social context” model and compared to final models described above. The same procedure was followed for mediation through type of accommodation.

Sensitivity analyses were carried out to check robustness of results, as listed in Supplementary File S3. Sensitivity analyses were carried out with relevant mediating variables as we were interested in the direct, not the total, effect of deprivation.

All analyses were conducted using Stata Version 15.

## Results

A higher proportion of the sample lived in the less deprived quintiles compared to the more deprived quintiles (Table 1). The sample is young, with and overall mean age of 35·0, and 60·2% of the sample is male. The largest share of participants was from Western Asia (63·6%), followed by South Asia (19·5%). The majority of participants entered the survey between 1-2 years after arrival in Germany (62·1%), with the remaining participants being approximately equally spread between having arrived under 1 year ago and over 2 years ago, while a small proportion (4·6%) arrived more than 3 years ago. On entry into the survey, half of participants (49·2%) had obtained a temporary refugee status in accordance with the Geneva convention, while 38·5% of participants were still waiting for the outcome of their asylum application. Other asylum status groups contributed less than 5% of participants to the sample. The state of North Rhine-Westphalia contributes the largest number of participants to the sample (40%), followed by Baden-Württemberg (21·2%), Bavaria (17·3%) and Hesse (12·3%), with the remaining states contributing less than 5% each. The distribution of the sample across federal states is markedly different by deprivation quintile. Socio-economic differences across deprivation quintiles are statistically significant for age, region of origin, months since arrival, asylum status and federal state. Differences in accommodation and social context variables across quintiles were statistically significant for type of accommodation, perception of neighbourhood safety, feeling socially isolated and the number of acquaintances from other countries of origin. Proportions and means for other indicators were comparable across quintiles (see Supplementary File S4).

**Table 1:** Socio-demographic characteristics of participants.

|  |  | Socio-Economic Deprivation (GISD) |  |  |  |  | Total | chi2-test<br>*p-value* |
| --- | --- | --- | --- | --- | --- | --- | --- | --- |
|  |  | Q1<br>(lowest) | Q2<br>(moderate-low) | Q3<br>(moderate) | Q4<br>(moderate-high) | Q5<br>(highest) |  |  |
| **Total sample** | n (%) | 470 (33·6%) | 420 (30·0%) | 217 (15·5%) | 182 (13·0%) | 111 (7·9%) | 1400<br>(100·0%) |  |
| **Gender** |  |  |  |  |  |  |  |  |
| male | n (%) | 291 (61·9%) | 256 (61·0%) | 130 (59·9%) | 111 (61·0%) | 55 (49·5%) | 843 (60·2%) | 0·200 |
| female | n (%) | 179 (38·1%) | 164 (39·0%) | 87 (40·1%) | 71 (39·0%) | 56 (50·5%) | 557 (39·8%) |  |
| *Total* | *n (%)* | *470 (100·0%)* | *420 (100·0%)* | *217 (100·0%)* | *182 (100·0%)* | *111 (100·0%)* | *1400*<br>*(100·0%)* |  |
| **Age** | Mean<br>(sd) | 33·9 (10·1) | 35·0 (10·2) | 36·1 (12·0) | 34·9 (11·4) | 37·5 (11·1) | 35·0 (10·7) | 0·012 |
| **Region of origin** |  |  |  |  |  |  |  |  |
| South Asia | n (%) | 107 (22·8%) | 85 (20·2%) | 29 (13·4%) | 24 (13·2%) | 28 (25·2%) | 273 (19·5%) | <0·001 |
| Southern Europe | n (%) | 13 (2·8%) | 4 (1·0%) | 4 (1·8%) | 2 (1·1%) | 1 (0·9%) | 24 (1·7%) |  |
| Western Asia | n (%) | 261 (55·5%) | 267 (63·6%) | 158 (72·8%) | 135 (74·2%) | 70 (63·1%) | 891 (63·6%) |  |
| East Africa | n (%) | 34 (7·2%) | 27 (6·4%) | 5 (2·3%) | 4 (2·2%) | 4 (3·6%) | 74 (5·3%) |  |
| West Africa | n (%) | 18 (3·8%) | 4 (1·0%) | 3 (1·4%) | 1 (0·5%) | 0 (0·0%) | 26 (1·9%) |  |
| Eastern Europe | n (%) | 7 (1·5%) | 4 (1·0%) | 5 (2·3%) | 0 (0·0%) | 2 (1·8%) | 18 (1·3%) |  |
| other/stateless | n (%) | 30 (6·4%) | 29 (6·9%) | 13 (6·0%) | 16 (8·8%) | 6 (5·4%) | 94 (6·7%) |  |
| *Total* | *n (%)* | *470 (100·0%)* | *420 (100·0%)* | *217 (100·0%)* | *182 (100·0%)* | *111 (100·0%)* | *1400*<br>*(100·0%)* |  |
| **Months since arrival** |  |  |  |  |  |  |  |  |
| 1-12 months | n (%) | 69 (15·7%) | 56 (14·3%) | 34 (16·9%) | 22 (12·4%) | 18 (17·8%) | 199 (15·2%) | 0·002 |
| 12-24 months | n (%) | 285 (64·9%) | 227 (57·9%) | 110 (54·7%) | 132 (74·6%) | 60 (59·4%) | 814 (62·1%) |  |
| >24 months | n (%) | 65 (14·8%) | 90 (23·0%) | 49 (24·4%) | 16 (9·0%) | 17 (16·8%) | 237 (18·1%) |  |
| >36 months | n (%) | 20 (4·6%) | 19 (4·8%) | 8 (4·0%) | 7 (4·0%) | 6 (5·9%) | 60 (4·6%) |  |
| *Total* | *n (%)* | *439 (100·0%)* | *392 (100·0%)* | *201 (100·0%)* | *177 (100·0%)* | *101 (100·0%)* | *1310*<br>*(100·0%)* |  |
| **Asylum status** |  |  |  |  |  |  |  |  |
| Asylum seeker (§55AsylG) | n (%) | 208 (44·5%) | 180 (43·5%) | 70 (32·6%) | 43 (24·2%) | 31 (28·4%) | 532 (38·5%) | <0·001 |
| Entitled to asylum (§25·1 AufenthG) | n (%) | 10 (2·1%) | 15 (3·6%) | 4 (1·9%) | 7 (3·9%) | 0 (0·0%) | 36 (2·6%) |  |
| Recognised refugee according to Geneva convention (§25·1 AufenthG) | n (%) | 201 (43·0%) | 181 (43·7%) | 118 (54·9%) | 118 (66·3%) | 63 (57·8%) | 681 (49·2%) |  |
| Unrestricted right to reside (§26·3 AufenthG) | n (%) | 3 (0·6%) | 0 (0·0%) | 1 (0·5%) | 0 (0·0%) | 1 (0·9%) | 5 (0·4%) |  |
| Temporary suspension of deportation (§60a AufenthG) | n (%) | 32 (6·9%) | 14 (3·4%) | 12 (5·6%) | 5 (2·8%) | 5 (4·6%) | 68 (4·9%) |  |
| Residence on humanitarian grounds | n (%) | 13 (2·8%) | 24 (5·8%) | 10 (4·7%) | 5 (2·8%) | 9 (8·3%) | 61 (4·4%) |  |
| *Total* | *n (%)* | *467 (100·0%)* | *414 (100·0%)* | *215 (100·0%)* | *178 (100·0%)* | *109 (100·0%)* | *1383*<br>*(100·0%)* |  |
| **Education** |  |  |  |  |  |  |  |  |
| primary education/<br>still in school | n (%) | 198 (45·0%) | 168 (42·1%) | 75 (36·6%) | 70 (42·4%) | 47 (43·5%) | 558 (42·4%) | 0·386 |
| lower secondary education | n (%) | 83 (18·9%) | 82 (20·6%) | 40 (19·5%) | 41 (24·8%) | 23 (21·3%) | 269 (20·4%) |  |
| Upper secondary education | n (%) | 70 (15·9%) | 78 (19·5%) | 46 (22·4%) | 26 (15·8%) | 17 (15·7%) | 237 (18·0%) |  |
| Post-secondary non-tertiary education | n (%) | 11 (2·5%) | 11 (2·8%) | 1 (0·5%) | 5 (3·0%) | 2 (1·9%) | 30 (2·3%) |  |
| Tertiary education | n (%) | 78 (17·7%) | 60 (15·0%) | 43 (21·0%) | 23 (13·9%) | 19 (17·6%) | 223 (16·9%) |  |
| *Total* | *n (%)* | *440 (100·0%)* | *399 (100·0%)* | *205 (100·0%)* | *165 (100·0%)* | *108 (100·0%)* | *1317*<br>*(100·0%)* |  |
| **federal state** |  |  |  |  |  |  |  |  |
| Northrhine-Westphalia | n (%) | 91 (19.4%) | 136 (32.4%) | 162 (74.7%) | 140 (76.9%) | 31 (27.9%) | 560 (40.0%) |  |
| Baden-Wuerttemberg | n (%) | 169 (36.0%) | 114 (27.1%) | 14 (6.5%) | 0 (0.0%) | 0 (0.0%) | 297 (21.2%) |  |
| Bavaria | n (%) | 142 (30.2%) | 64 (15.2%) | 23 (10.6%) | 17 (9.3%) | 0 (0.0%) | 246 (17.6%) |  |
| Hessia | n (%) | 68 (14.5%) | 89 (21.2%) | 15 (6.9%) | 0 (0.0%) | 0 (0.0%) | 172 (12.3%) |  |
| Saarland | n (%) | 0 (0.0%) | 10 (2.4%) | 2 (0.9%) | 8 (4.4%) | 0 (0.0%) | 20 (1.4%) |  |
| Saxony | n (%) | 0 (0.0%) | 7 (1.7%) | 1 (0.5%) | 17 (9.3%) | 19 (17.1%) | 44 (3.1%) | <0.001 |
| Saxony-Anhalt | n (%) | 0 (0.0%) | 0 (0.0%) | 0 (0.0%) | 0 (0.0%) | 61 (55.0%) | 61 (4.4%) |  |
| Total | n (%) | 470 (100.0%) | 420 (100.0%) | 217 (100.0%) | 182 (100.0%) | 111 (100.0%) | 1400 (100.0%) |  |

The sample covers 485 communes, out of a total 11 116 communes in Germany. While the lowest deprivation quintiles are covered with a substantial number of communes, participants in the higher deprivation quintiles are concentrated in fewer communes (Supplementary File S5).

Descriptively, a small improvement in mcs score (0·3, sd: 14·3) can be observed between t0 and t1 across the sample, but there is a substantial improvement in the highest deprivation quintile (Figure 2). With regard to the pcs score, there is a small decline (-0·6, sd: 10·4) between t0 and t1 across the sample, but no discernable pattern can be observed between quintiles (see also Supplementary File S6).

**Figure 2:**
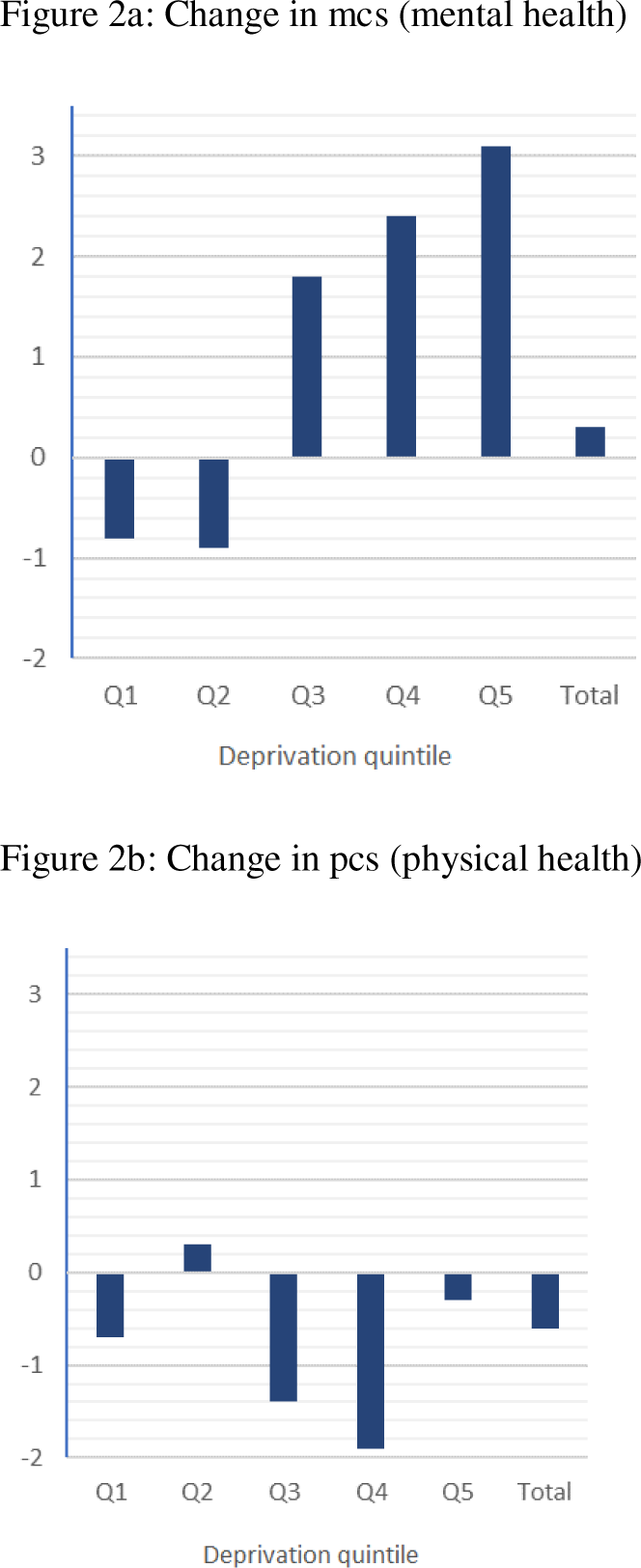
Change in mcs (mental health) and pcs (physical health) between baseline and follow-up by deprivation quintile. Q1: lowest deprivation; Q2: moderate-low deprivation; Q3: moderate deprivation; Q4: moderate-high deprivation; Q5: highest deprivation

Single-level linear regression models show an improvement in mental health (mcs score) in the most deprived quintiles, with consistent positive coefficients for Q4 (moderate-high deprivation) and Q5 (highest deprivation) compared to Q1 (lowest deprivation) across crude, baseline and sociodemographic models (Table 2). However, differences are leveled to statistical non-significance once baseline and socio-demographic characteristics have been adjusted for. Model fit is acceptable, with a large improvement of adjusted r2 (ar2) in the baseline model (ar2=0·40) and some improvement in the socio-demographic model (ar2=0·42) compared to the crude model (ar2=0·008). Wald tests show that introduction of baseline variables is significant (p<0·001) compared to the null model and the introduction of sociodemographic variables significant (p=0·04) compared to the baseline model. When applying a multi-level model to the mental health outcome, the null model provides evidence of clustering at community level (ICC: 0·095; LRtest p<0·001). The final multi-level model, adjusted for baseline and socio-demographic variables, confirms the results of the linear models: individuals living in areas of moderate-high deprivation (Q4: coef·: 2·5, 95%CI: -0·7;5·6) and highest deprivation (Q5: coef·: 2·1, 95%CI: - 3·4;7·7) show improvements in mental health compared to individuals living in areas of lowest deprivation (Q1), but these are not statistically significant (Table 2).

**Table 2:** Results of single-level and multi-level regression models for mental (mcs) and physical (pcs) health outcomes. Mcs= mental health component summary scale; pcs= physical health component summary scale Significance levels: + p<0·1; *p<0·05; **p<0·01

|  | Mcs (t0-t1) |  |  |  | Pcs (t0-t1) |  |  |  |
| --- | --- | --- | --- | --- | --- | --- | --- | --- |
|  | crude | baseline | sociodemo | Multi-level | crude | baseline | sociodemo | Multi-level |
| Clustering | None | None | None | commune | None | None | None | commune |
| Q1 (least deprived) | ref | ref | ref | ref | ref | ref | ref | ref |
| Q2 | -0.07073<br>(-2.00746 - 1.86600) | 0.67528<br>(-0.90816 - 2.25873) | 0.29488<br>(-1.40286 - 1.99261) | **0.13598**<br>**(-1.50919 - 1.78115)** | 0.95734<br>(-0.36240 - 2.27709) | 0.20278<br>(-0.98853 - 1.39409) | **-0.10757**<br>**(-1.33126 - 1.11613)** | -0.09651<br>(-1.27228 - 1.07926) |
| Q3 | 2.54863*<br>(0.37301 - 4.72424) | 1.04715<br>(-0.88023 - 2.97453) | 0.49263<br>(-1.60776 - 2.59302) | **0.15890**<br>**(-1.99852 - 2.31632)** | -0.71674<br>(-2.26403 - 0.83055) | -0.75427<br>(-2.26029 - 0.75175) | **-0.90339**<br>**(-2.43377 - 0.62699)** | -0.86688<br>(-2.47375 - 0.73998) |
| Q4 | 3.21475*<br>(0.60830 - 5.82119) | 3.13700**<br>(0.88526 - 5.38875) | 2.22299+<br>(-0.16188 - 4.60786) | **2.45583**<br>**(-0.72454 - 5.63620)** | -1.20706<br>(-3.13981 - 0.72569) | -1.62063+<br>(-3.53884 - 0.29758) | **-2.15155\***<br>**(-4.10117 - -0.20192)** | -2.12254+<br>(-4.25471 - 0.00962) |
| Q5 | 3.90317**<br>(1.19714 - 6.60921) | 3.05355<br>(-0.81873 - 6.92584) | 2.16679<br>(-2.11464 - 6.44821) | **2.11347**<br>**(-3.44231 - 7.66925)** | 0.35001<br>(-1.78872 - 2.48874) | 0.27058<br>(-2.73707 - 3.27824) | **-0.97769**<br>**(-4.22444 - 2.26907)** | -0.93641<br>(-4.86835 - 2.99553) |
| Mcs/pcs t0 |  | -0.75852**<br>(-0.81325 - -0.70378) | -0.77411**<br>(-0.83182 - -0.71640) | **-0.76848\*\***<br>**(-0.82448 - -0.71248)** |  | -0.53983**<br>(-0.59344 - -0.48622) | **-0.67844\*\***<br>**(-0.73829 - -0.61859)** | -0.67846**<br>(-0.73947 - -0.61745) |
| South Asia |  | ref | ref | ref |  | ref | ref | ref |
| Southern Europe |  | -0.68440<br>(-6.01609 - 4.64729) | -1.09137<br>(-7.78360 - 5.60086) | **-0.65495**<br>**(-7.07214 - 5.76224)** |  | -1.22632<br>(-4.66457 - 2.21194) | **-1.72344**<br>**(-6.05972 - 2.61283)** | -1.73537<br>(-6.34731 - 2.87658) |
| Western Asia |  | 2.63345**<br>(1.00534 - 4.26157) | 2.63402**<br>(0.83974 - 4.42830) | **2.50945\*\***<br>**(0.66031 - 4.35859)** |  | -1.57268*<br>(-2.87898 - -0.26638) | **-0.71329**<br>**(-2.20108 - 0.77450)** | -0.70737<br>(-2.20916 - 0.79441) |
| East Africa |  | 4.72106**<br>(2.00496 - 7.43715) | 5.34769**<br>(2.42157 - 8.27381) | **5.37214\*\***<br>**(2.45207 - 8.29221)** |  | 0.35500<br>(-1.78905 - 2.49906) | **0.45291**<br>**(-1.48183 - 2.38765)** | 0.45821<br>(-1.51923 - 2.43565) |
| West Africa |  | 4.19238+<br>(-0.33078 - 8.71555) | 5.26274*<br>(0.39699 - 10.12850) | **5.14355+**<br>**(-0.14284 - 10.42994)** |  | -1.98694<br>(-5.39318 - 1.41930) | **-2.09123**<br>**(-6.03411 - 1.85165)** | -2.07402<br>(-6.15570 - 2.00766) |
| Eastern Europe |  | 4.49096<br>(-1.06378 - 10.04570) | 5.54476+<br>(-0.09088 - 11.18039) | **5.74738\***<br>**(0.26748 - 11.22729)** |  | -1.13899<br>(-5.04487 - 2.76689) | **-0.17281**<br>**(-4.80573 - 4.46010)** | -0.18125<br>(-4.38673 - 4.02423) |
| Other/stateless |  | 1.51103<br>(-1.26953 - 4.29159) | 2.17797<br>(-0.61300 - 4.96894) | **2.12388**<br>**(-0.85449 - 5.10224)** |  | -1.18163<br>(-3.25296 - 0.88970) | **-0.78043**<br>**(-3.17270 - 1.61183)** | -0.78065<br>(-3.18510 - 1.62380) |
| Northrhine-Westphalia |  | ref | ref | ref |  | ref | ref | ref |
| Hessia |  | 0.26165<br>(-1.90214 - 2.42544) | -0.76118<br>(-3.16125 - 1.63890) | **-0.40102**<br>**(-3.02785 - 2.22582)** |  | 0.65813<br>(-1.02118 - 2.33744) | **1.48846**<br>**(-0.34505 - 3.32198)** | 1.48504<br>(-0.35364 - 3.32373) |
| Baden-Wuerttemberg |  | -1.83822*<br>(-3.60120 - -0.07524) | -2.53865**<br>(-4.46921 - -0.60808) | **-2.51632\***<br>**(-4.49104 - -0.54159)** |  | 0.06681<br>(-1.29055 - 1.42416) | **0.34651**<br>**(-1.10152 - 1.79455)** | 0.35395<br>(-1.07480 - 1.78271) |
| Bavaria |  | 0.83086<br>(-1.07960 - 2.74133) | 0.06519<br>(-1.87479 - 2.00516) | **0.16860**<br>**(-1.97459 - 2.31178)** |  | 0.20124<br>(-1.24860 - 1.65108) | **0.30255**<br>**(-1.21099 - 1.81610)** | 0.30496<br>(-1.18567 - 1.79558) |
| Saarland |  | -1.92863<br>(-5.78780 - 1.93055) | -2.42567<br>(-6.88264 - 2.03129) | **-2.58823**<br>**(-6.71085 - 1.53439)** |  | 2.03714<br>(-2.01982 - 6.09410) | **2.15096**<br>**(-1.63435 - 5.93627)** | 2.15350<br>(-1.15803 - 5.46503) |
| Saxony |  | -1.49535<br>(-5.32045 - 2.32974) | -0.13311<br>(-3.78322 - 3.51700) | **0.30401**<br>**(-3.55018 - 4.15820)** |  | -1.99098<br>(-5.34330 - 1.36134) | **0.03143**<br>**(-3.35589 - 3.41875)** | 0.02298<br>(-3.23220 - 3.27816) |
| Saxony-Anhalt |  | -0.34989<br>(-4.36212 - 3.66234) | -0.37396<br>(-4.80999 - 4.06208) | **0.07399**<br>**(-5.45825 - 5.60623)** |  | -0.58409<br>(-4.11143 - 2.94325) | **2.22915**<br>**(-1.52027 - 5.97856)** | 2.18273<br>(-2.06836 - 6.43382) |
| 1-12 months |  |  | ref | ref |  |  | ref | ref |
| 12-24 months |  |  | -0.68779<br>(-2.44904 - 1.07346) | **-0.53589**<br>**(-2.32725 - 1.25547)** |  |  | **0.52219**<br>**(-0.76068 - 1.80505)** | 0.51293<br>(-0.82702 - 1.85287) |
| 24-36 months |  |  | 1.03611<br>(-1.09716 - 3.16938) | **1.04139**<br>**(-1.12793 - 3.21071)** |  |  | **1.05831**<br>**(-0.53580 - 2.65242)** | 1.06198<br>(-0.59042 - 2.71438) |
| >36 months |  |  | -2.39996<br>(-5.84250 - 1.04258) | **-2.52655**<br>**(-6.25308 - 1.19999)** |  |  | **1.05658**<br>**(-1.63487 - 3.74803)** | 1.04662<br>(-1.67204 - 3.76527) |
| male |  |  | ref | ref |  |  | ref | ref |
| female |  |  | -1.43967* | **-1.35709\*** |  |  | **-3.21420\*\*** | -3.21612** |
|  |  |  | (-2.75896 - -0.12038) | (-2.67589 - -0.03828) |  |  | (-4.25138 - -2.17701) | (-4.26321 - -2.16902) |
| primary education/ still in school |  |  | ref | ref |  |  | ref | ref |
| lower secondary education |  |  | 1.63739*<br>(0.00428 - 3.27050) | 1.44832+<br>(-0.20932 - 3.10595) |  |  | -0.54066<br>(-1.87264 - 0.79132) | -0.53709<br>(-1.94286 - 0.86869) |
| Upper secondary education |  |  | -0.04855<br>(-1.81639 - 1.71929) | -0.07931<br>(-1.83225 - 1.67364) |  |  | 0.75682<br>(-0.56230 - 2.07593) | 0.76190<br>(-0.58932 - 2.11312) |
| Post-secondary non-tertiary education |  |  | -2.65972<br>(-6.56836 - 1.24893) | -2.22532<br>(-6.23514 - 1.78450) |  |  | 0.45026<br>(-2.79452 - 3.69504) | 0.41506<br>(-2.79714 - 3.62726) |
| Tertiary education |  |  | -0.30467<br>(-2.17099 - 1.56165) | -0.24362<br>(-2.18526 - 1.69802) |  |  | 2.06805**<br>(0.71749 - 3.41860) | 2.06576**<br>(0.73200 - 3.39952) |
| age |  |  | -0.01798<br>(-0.08407 - 0.04811) | -0.01874<br>(-0.08374 - 0.04627) |  |  | -0.23104**<br>(-0.28385 - -0.17824) | -0.23100**<br>(-0.28329 - -0.17872) |
| Asylum seeker (§55AsylG) |  |  | ref | ref |  |  | ref | ref |
| Entitled to asylum (§25.1 AufenthG) |  |  | 2.44898<br>(-1.15197 - 6.04992) | 2.50640<br>(-0.97058 - 5.98338) |  |  | 2.93585*<br>(0.60565 - 5.26606) | 2.92380*<br>(0.33651 - 5.51110) |
| Recognised refugee according to Geneva convention (§25.1 AufenthG) |  |  | 0.83330<br>(-0.81628 - 2.48288) | 1.01473<br>(-0.63460 - 2.66405) |  |  | 0.37400<br>(-0.89789 - 1.64589) | 0.35923<br>(-0.91231 - 1.63078) |
| Unrestricted right to reside (§26.3 AufenthG) |  |  | -4.44847<br>(-12.72726 - 3.83032) | -4.37722<br>(-12.57098 - 3.81654) |  |  | -3.66379<br>(-13.11546 - 5.78788) | -3.68614<br>(-13.61946 - 6.24718) |
| Temporary suspension of deportation (§60a AufenthG) |  |  | -1.70100<br>(-5.58338 - 2.18139) | -1.86562<br>(-5.84480 - 2.11355) |  |  | 1.91872<br>(-0.56349 - 4.40093) | 1.90518<br>(-0.68586 - 4.49623) |
| Residence on humanitarian grounds |  |  | 0.09225<br>(-3.18342 - 3.36792) | 0.30565<br>(-2.93621 - 3.54752) |  |  | 0.45074<br>(-2.34604 - 3.24751) | 0.44840<br>(-2.26599 - 3.16279) |
| Observations | 1400 | 1400 | 1226 | 1226 | 1400 | 1400 | 1226 | 1226 |
| R-squared | 0.01148 | 0.41053 | 0.43523 |  | 0.00509 | 0.28350 | .3655612 |  |
| adjusted R-squared | 0.00865 | 0.40328 | 0.42057 |  | 0.00224 | 0.27468 | .3490891 |  |
| model degrees of freedom | 4 | 17 | 31 | 31 | 4 | 17 | 31 | 31 |
| Number of clusters |  |  |  | 439 |  |  |  | 439 |
| Aikikes information criterion (AIC) |  |  |  | 9369.444 |  |  |  | 8757.678 |
| Bayes information criterion (BIC) |  |  |  | 9543.236 |  |  |  | 8931.47 |
| Intra-class correlation coefficient (ICC) |  |  |  | 0.07884 |  |  |  | .0032675 |
| Neighbourhood intercept variance |  |  |  | 9.22415 |  |  |  | 0.22906 |
| Residual variance |  |  |  | 107.7828 |  |  |  | 69.87543 |
| Proportional change in variance vs. null model (PCV<sub>N</sub>) |  |  |  | 53% |  |  |  | 87% |
| Likelihood-ratio test vs. linear model |  |  |  | p=0.0004 |  |  |  | p=0.4148 |

For physical health (pcs score), single-level regression models show a dose response relationship, with individuals in quintiles with higher deprivation reporting worse physical health (Table 2). The strength of this relationship increases as baseline and sociodemographic variables are introduced. However, only the decline in physical health for individuals living in areas of moderate-high deprivation (Q4) compared to lowest deprivation (Q1) is statistically significant once socio-demographic characteristics have been adjusted for (Q4: coef·: 2·2, 95%CI: -4·1;-0·2). Model fit is acceptable, with a large improvement of adjusted r2 in baseline model (0·27) and some improvement in socio-demographic model (0·35) compared to crude model (0·002), but some variation is still unaccounted for. Wald tests show that the introduction of baseline variables is significant (p<0·001) compared to the null model and the introduction of sociodemographic variables is significant (p<0·001) compared to the baseline model. Applying multi-level modelling to the physical health outcome, the null model does not suggest that clustering is occurring at the community level (ICC: 0·01; LRtest p=0·103). Fully adjusted multi-level models shows near-identical results to the simple linear regression (Table 2). Given these results, multi-level modelling will be applied for mental health, but not physical health models. VIF were small for all variables included in the final mental and physical models (see Supplementary File S7).

Considering potential mediation variables, a statistically significant association can be observed between the mental health outcome and subjective feelings of social isolation, feeling like an outsider, feeling welcome and worries about xenophobia, as well as a borderline significance for the number of acquaintances from the country of origin (see Supplementary File S8). No relationship can be observed between the social context variables and the physical health outcome. The type of accommodation showed no discernible descriptive relationship with change in either mental or physical health score (see Supplementary File S8). In regression-based mediation analyses, one-by-one introduction of social context variables in the mental health model (multi-level) revealed that the following variables showed an improvement in model fit: safety of the neighbourhood (p=0·01), feeling socially isolated (p=0·0001), feeling like an outsider (p=0·008), worries about xenophobia (p<0·0001). Including these social context variables in the model reduced the positive effect of moderate-high deprivation on mental health compared to the fully adjusted model (Q4: coef·: 2·2, 95%CI:-0·6;5·0), with no statistically significant differences between quintiles (Figure 2a, see also Supplementary File S9)). Introducing social context variables into the physical health model had a negligible effect on the relationship between deprivation and physical health; a dose-response relationship with the negative impact of deprivation can still be observed which is statistically significant for moderate-high deprivation (Q4: coef·:-2·4, 95%CI:-4·3;-0·5; Figure 2b, see also Supplementary File S8). Type of accommodation did not show a statistically significant improvement in model fit for either health outcome (see Supplementary File S8).

**Figure 3:**
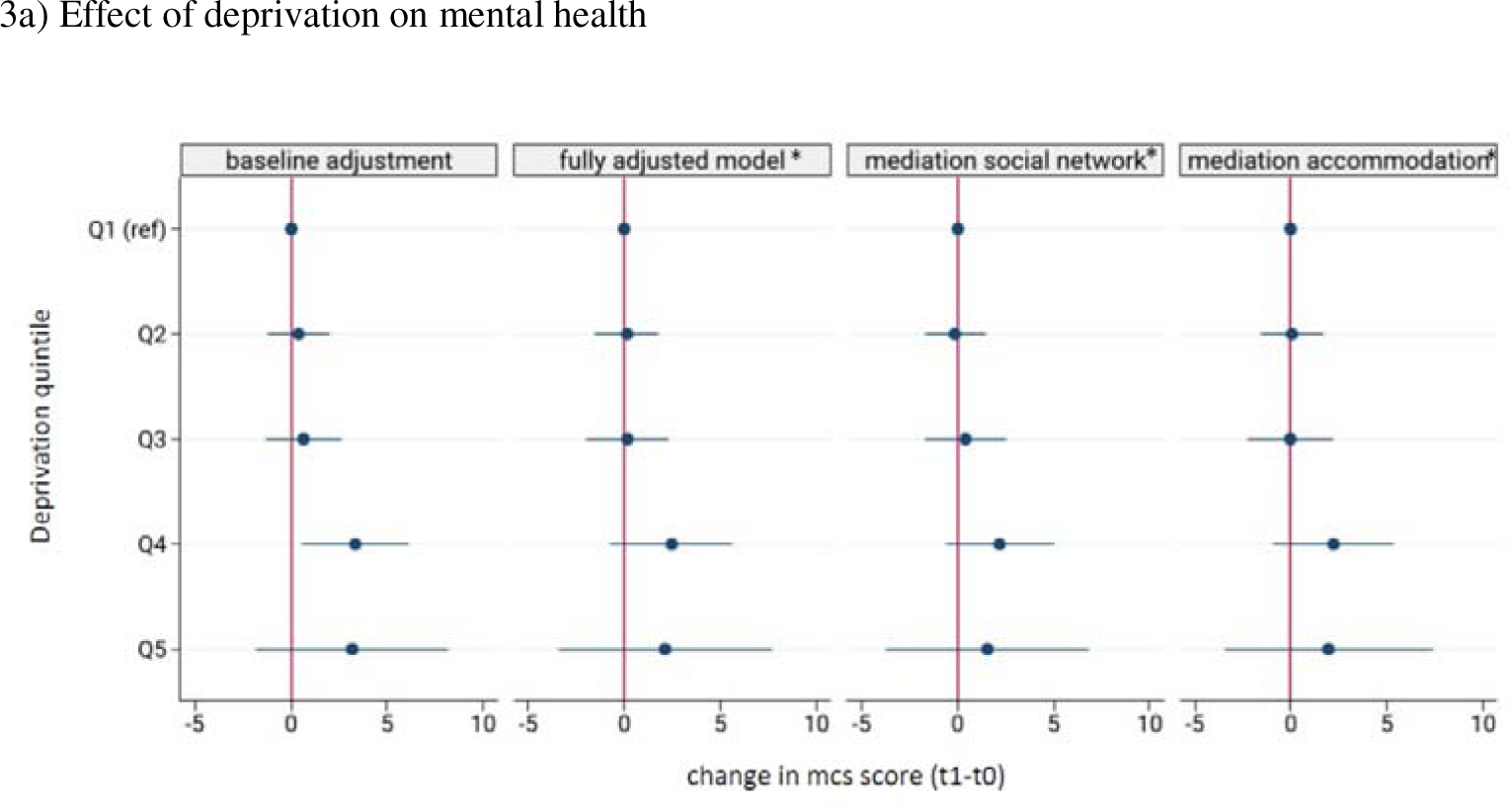
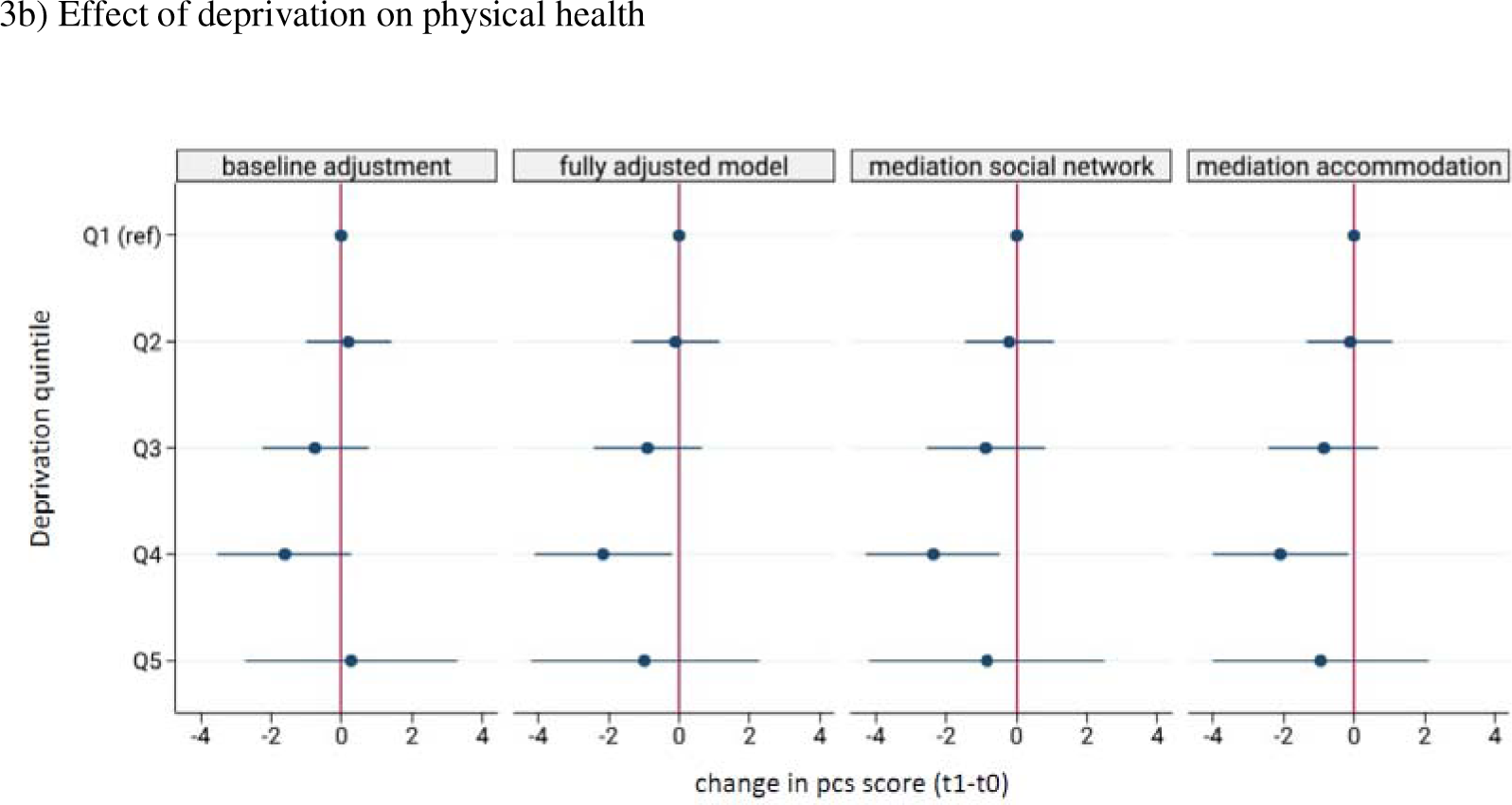
Coefficient plots for baseline and fully adjusted models as well as mediation analyses. * Multi-level model accounting for random slopes at the commune level mcs= mental health component scale of the SF-12; pcs= physical health component scale of the SF-12

Sensitivity analyses adjusting by years of follow-up and wave, excluding those who are not state assigned at t1, excluding outliers and applying inverse probability weighting (S1-S5& S7) had only negligible effects on the main results of the mental health model. When the sample was determined by subjectively assessed residential assignment, however, differences in mcs between the least deprived and other quintiles were small, and none were statistically significant (see Supplementary File S10, S12 & S13). Results for the physical health outcome remained stable through all sensitivity analyses: the dose-response relationship continued to be evident and was statistically significant for Q4 vs. Q1 in all but one sensitivity analysis (S4: excluding outliers) (see Supplementary File S11, S12 & S13).

## Discussion

This study demonstrates the negative impact of small-area deprivation on the physical health of residents. This result is robust to sensitivity analyses and not mediated by accommodation or social context variables. Furthermore, there is some evidence of a positive impact of deprivation on mental health, some of which may be explained through social context, although this effect is statistically insignificant and not entirely robust to sensitivity analysis.

The results of this study confirm the existing evidence ^1^, including other natural experiments ^4,18,19^, on the relationship between deprivation and physical health. In contrast to previous studies, which considered the effects over long time periods, our analysis shows that the negative effects of deprivation are evident even after 1-2 years of follow-up. Deprivation may have cumulative, long-term effects on the physical health status of residents through diminished neighbourhood resources such as access to green space, food availability, walkability and environmental pollution. However, our analysis suggests that availability and accessibility of health and social care structures, which have more immediate effects, play an important role in determining refugee health in more deprived regions. While recognised refugees are integrated into regular healthcare and social service infrastructure, important access barriers, including language skills, a lack of awareness of services and difficulties navigating complex bureaucratic systems have been documented ^20^. Adequate provision of interpreting services, support navigating systems through social workers and accessible information is therefore crucial in supporting accessibility and ensuring adequate coverage of services. Areas with higher deprivation may have less expendable resources to invest in services and their accessibility for refugees. In order to avoid further deterioration of health in the more deprived regions, the delivery of health and social services in these regions should be supported at the level of the federal states, for example by using deprivation as a criterion in the allocation of available integration budgets.

The results of our study further contribute to the growing body of literature which shows complex effects of deprivation on mental health. While some studies have reported a negative impact of deprivation on symptoms of depression and anxiety ^21,22^, our study and other natural experiments using strict dispersal policies among refugees ^23^ suggest that this may be due to selection effects. Robust evidence from other natural experiments does, however, provide support for the negative effects of deprivation on psychiatric diagnoses and prescriptions^24,25^.

In fact, our study shows suggestive evidence of a positive impact of regional deprivation on mental health. A possible explanation for this effect is the relative income hypothesis ^26^, which suggests that the *relative* inequality experienced by refugees residing in areas of low deprivation might incur higher levels of stress compared to those living in areas with higher deprivation, where wealth and status differentials between the resident population and newly arriving refugees are less pronounced. However, a natural experiment assessing the effect of income inequality on health among refugees found no relationship with mental health outcomes ^27^. The effect may also be explained by the social context in more deprived regions if social attributes of these areas are supportive for mental health ^22^. The effects of social context variables in this analysis were small. However, we only considered individual-level social context attributes. Further mechanisms should be explored, including a consideration of area-level attributes related to social cohesion, including aspects such as voting behaviour, civic participation, racist incidents and migrant density. Co-ethnic migrant networks, in particular, may be more frequent in areas of high deprivation and could have protective effects by reducing assimilation pressures.

Strengths of this study are the natural experiment design with robust, multi-level difference-in-difference analysis which allows for causal interpretations. The study further benefits from robust survey data which allows for the direct identification of refugees as well as analysis of potential causal pathways. However, the causal interpretation of results is limited by the short follow-up time of 1-2 years and relatively small sample size, especially in the more deprived quintiles. Combined with the fact that surveys take place relatively soon after arrival in Germany, this means that changes in mental health status may reflect secular trends rather than responses to contextual characteristics. Further research with larger sample sizes and longer follow-up is required to confirm our results. A larger sample size would also allow for the use of nationality rather than region of origin as a covariate, which is key to adjust for important differences both in the dispersal process and pre-migration experiences. While the study uses small-area level contextual variables, these constitute political boundaries which may or may not refugees’ actual experienced communities ^3^. Alternative approaches which centres individuals’ experiences of their communities, such as social network modelling, is encouraged.

## Conclusions

This study finds a negative impact of deprivation on the physical health of refugees in Germany. Given the short timeframe of analysis, this can be attributed to gaps in health and social service provision. Further efforts should be made to support resource-poor regions to improve integration of refugees into health and social systems, including improved interpreting services, specifically trained social workers and diversity-sensitive information offerings. This should include an acknowledgement of already existing, informal social support networks among migrants, for example through the use of participatory action and peer education. Further research using longer timeframes and larger sample sizes are required to confirm the results of this study. These should further explore the mechanisms through which deprivation acts on health, in particular relating to the complex relationship with mental health, in order to guide future policy efforts.

## Data Availability

Data of the Socioeconomic Panel is publicly available for individuals at research institutions subject to eligibility requirements and data sharing agreements. Requests can be made via.

## Contributors

LB: Conceptualization, Methodology, Investigation, Formal analysis, Writing – Original Draft, Visualization, Project administration

KB: Conceptualization, Methodology, Writing – Review & Editing, Supervision, Funding acquisition

## Acknowledgements

This study was funded by the German Science Foundation (DFG) in the scope of the NEXUS project as part of the PH-LENS Research Unit (FOR 2928 / GZ: BO 5233/1-1). The funder had no influence on the design of the study, analysis or decision to publish.

## Supplementary File

### Text S1: Additional details of the refugee dispersal process in Germany

Upon arrival in Germany, refugees are dispersed into communes based on a three-level dispersal process at federal, regional and communal levels. First, refugees are assigned to one of Germany’s 16 federal states based on an administrative quota (“Königsteiner Schlüssel”) based on population size and tax revenue ^1^. In this process, the nationality of refugees is taken into account, since different regional offices of the Federal Ministry for Migration and refugees (BAMF) have specialisations for processing asylum seekers from different countries of origin ^2^.

The process for dispersal *within* federal states differs from state to state. After the asylum claim is formally lodged, some states disperse asylum seekers on to further reception centres within the state (second-level dispersal) before transferring them to communes (third-level dispersal), while others disperse refugees directly to communes. Refugees may be housed in state reception centres for up to 18 months, or until the end of the asylum process for refugees from so-called “safe” countries of origin. Some federal states have specially designated accommodation facilities for single women, families, or refugees with special needs or health issues. An exception to this are so-called “family reunification” refugees, which are family members of recognised refugees who are permitted to reside with their families for the duration of their asylum application. Here, selection of residence is endogenous given as no dispersal process is involved and individuals are not subject to mobility restrictions.

A further feature of the asylum system in Germany which makes it a unique natural experiment is the residence requirement policy (“Wohnsitzauflage”). The policy requires asylum seekers to remain resident in the region to which they were assigned for the duration of their asylum application and up to 3 years following a decision. The policy is only enacted in several federal states and applies to the following individuals: those who have not yet received a decision on their asylum decision, those who have received a negative asylum decision but have not yet left the country, those who have received a negative asylum decision but have been granted a temporary right to remain and those who have a positive asylum decision but are dependent on state benefits for a period of 3 years following the asylum decision ^3^. The latter category was included as an extension to the prior residential policy in several federal states from 2016 to 2018, but the timing of this policy change was not uniform ^4^.

---

**Figure S2:**
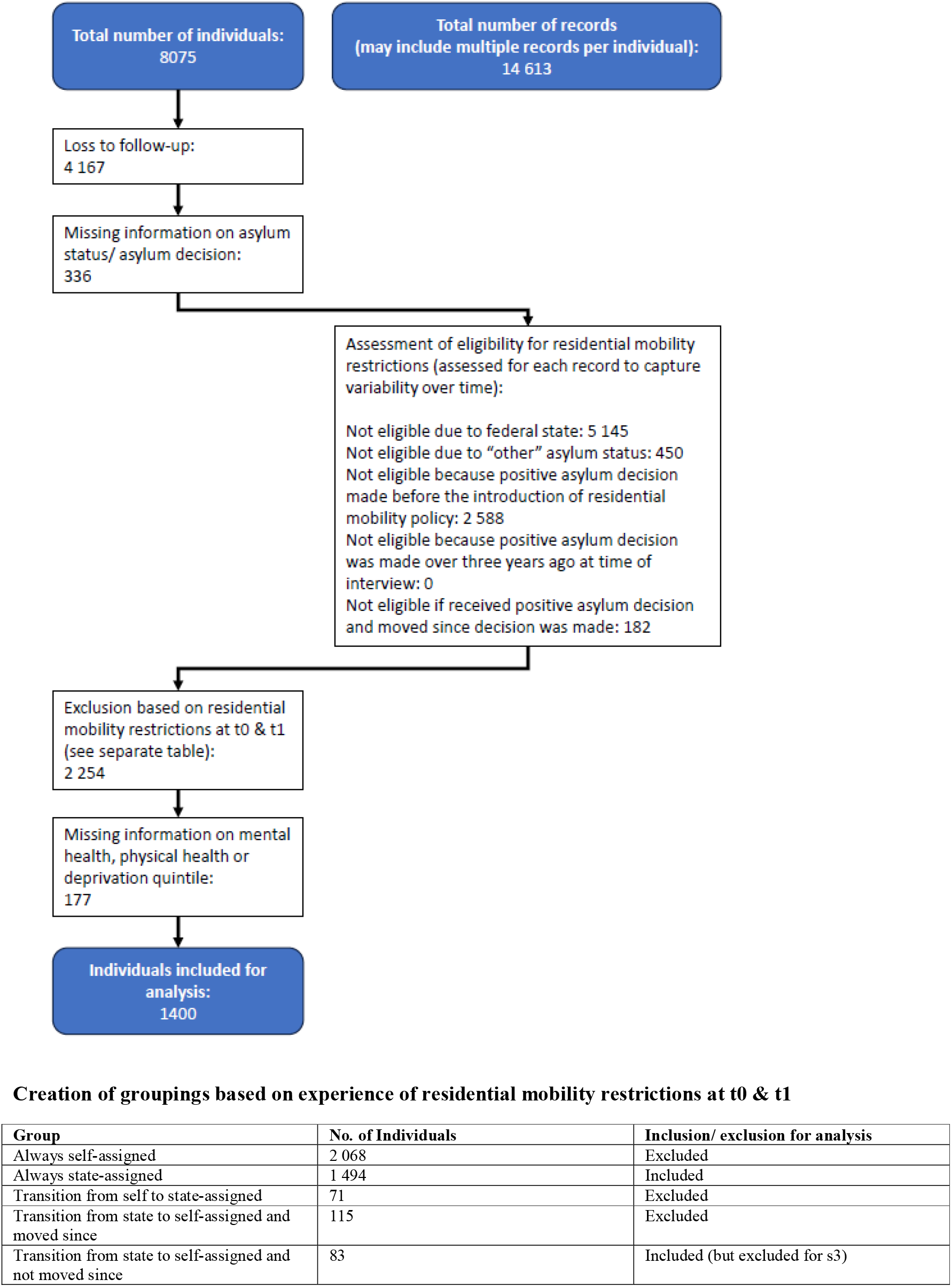
Flowchart of sample selection.

**Table S3:**
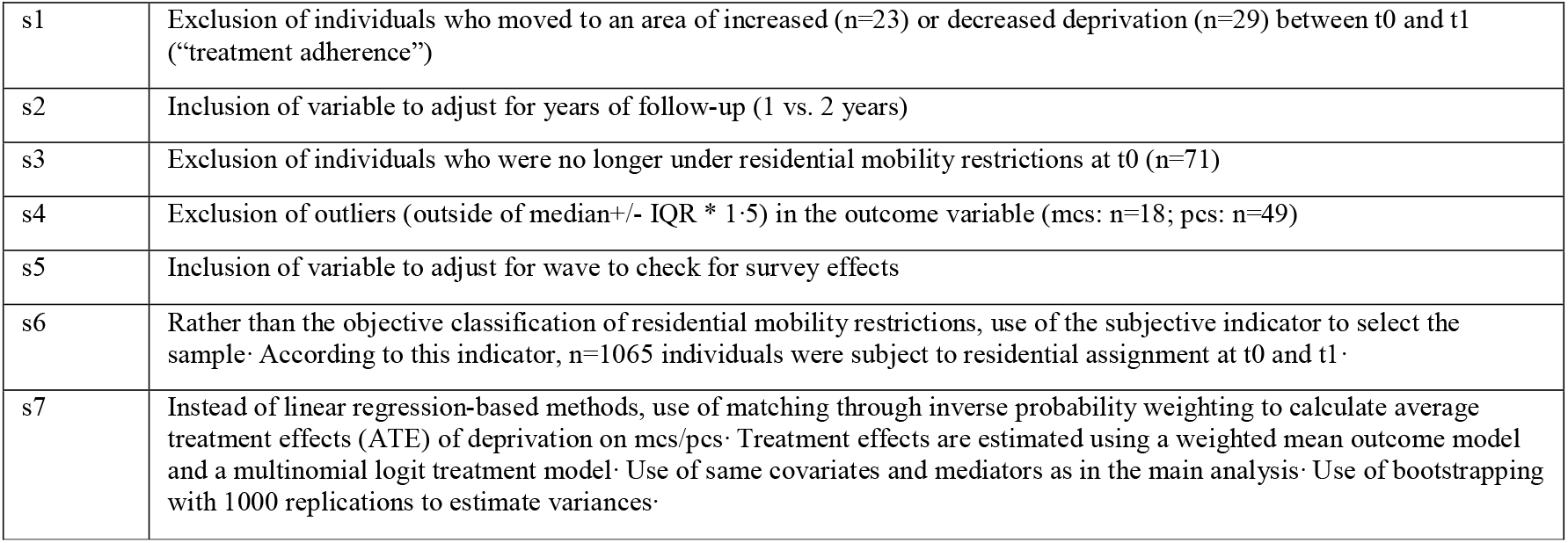
Additional details of sensitivity analyses.

**Table S4:** accommodation, social context and sample characteristics of study participants.

|  | Q1<br>(lowest) | Q2<br>(moderate-low) | Q3<br>(moderate) | Q4<br>(moderate-high) | Q5<br>(highest) | Total | Chi2-test<br>p-value |
| --- | --- | --- | --- | --- | --- | --- | --- |
| type of accommodation |  |  |  |  |  |  |  |
| Shared | 269 (57.4%) | 171 (40.9%) | 60 (27.6%) | 37 (20.3%) | 19 (17.1%) | 556 (39.8%) | <0.001 |
| Private | 200 (42.6%) | 247 (59.1%) | 157 (72.4%) | 145 (79.7%) | 92 (82.9%) | 841 (60.2%) |  |
| Total | 469 (100.0%) | 418 (100.0%) | 217 (100.0%) | 182 (100.0%) | 111 (100.0%) | 1397 (100.0%) |  |
| perception of neighborhood safety |  |  |  |  |  |  |  |
| Very safe | 366 (77.9%) | 369 (87.9%) | 164 (75.6%) | 148 (81.3%) | 82 (73.9%) | 1129 (80.6%) | <0.001 |
| Fairly safe | 80 (17.0%) | 40 (9.5%) | 49 (22.6%) | 26 (14.3%) | 26 (23.4%) | 221 (15.8%) |  |
| Fairly/ very unsafe | 24 (5.1%) | 11 (2.6%) | 4 (1.8%) | 8 (4.4%) | 3 (2.7%) | 50 (3.6%) |  |
| Total | 470 (100.0%) | 420 (100.0%) | 217 (100.0%) | 182 (100.0%) | 111 (100.0%) | 1400 (100.0%) |  |
| Feeling socially isolated |  |  |  |  |  |  |  |
| Very often | 39 (8.4%) | 36 (8.8%) | 15 (7.0%) | 14 (7.7%) | 3 (2.7%) | 107 (7.8%) | 0.003 |
| Often | 53 (11.4%) | 45 (10.9%) | 38 (17.8%) | 14 (7.7%) | 24 (21.8%) | 174 (12.6%) |  |
| Sometimes | 119 (25.7%) | 81 (19.7%) | 45 (21.0%) | 44 (24.3%) | 26 (23.6%) | 315 (22.8%) |  |
| Seldom | 89 (19.2%) | 68 (16.5%) | 43 (20.1%) | 26 (14.4%) | 17 (15.5%) | 243 (17.6%) |  |
| Never | 163 (35.2%) | 181 (44.0%) | 73 (34.1%) | 83 (45.9%) | 40 (36.4%) | 540 (39.2%) |  |
| Total | 463 (100.0%) | 411 (100.0%) | 214 (100.0%) | 181 (100.0%) | 110 (100.0%) | 1379 (100.0%) |  |
| Feeling like an outsider |  |  |  |  |  |  |  |
| Very often | 36 (7.9%) | 35 (8.6%) | 21 (10.0%) | 20 (11.2%) | 6 (5.5%) | 118 (8.6%) | 0.217 |
| Often | 60 (13.1%) | 49 (12.0%) | 33 (15.6%) | 18 (10.1%) | 21 (19.1%) | 181 (13.3%) |  |
| Sometimes | 136 (29.7%) | 100 (24.6%) | 50 (23.7%) | 52 (29.1%) | 35 (31.8%) | 373 (27.3%) |  |
| Seldom | 91 (19.9%) | 85 (20.9%) | 46 (21.8%) | 26 (14.5%) | 19 (17.3%) | 267 (19.6%) |  |
| Never | 135 (29.5%) | 138 (33.9%) | 61 (28.9%) | 63 (35.2%) | 29 (26.4%) | 426 (31.2%) |  |
| Total | 458 (100.0%) | 407 (100.0%) | 211 (100.0%) | 179 (100.0%) | 110 (100.0%) | 1365 (100.0%) |  |
| Feeling welcome |  |  |  |  |  |  |  |
| Completely | 291 (62.0%) | 269 (64.4%) | 122 (56.7%) | 132 (72.5%) | 71 (64.5%) | 885 (63.5%) | 0.161 |
| For the most part | 98 (20.9%) | 78 (18.7%) | 54 (25.1%) | 25 (13.7%) | 17 (15.5%) | 272 (19.5%) |  |
| In some respects | 58 (12.4%) | 50 (12.0%) | 23 (10.7%) | 18 (9.9%) | 15 (13.6%) | 164 (11.8%) |  |
| Hardly/ not at all | 22 (4.7%) | 21 (5.0%) | 16 (7.4%) | 7 (3.8%) | 7 (6.4%) | 73 (5.2%) |  |
| Total | 469 (100.0%) | 418 (100.0%) | 215 (100.0%) | 182 (100.0%) | 110 (100.0%) | 1394 (100.0%) |  |
| Worries about hostility to foreigners |  |  |  |  |  |  |  |
| A lot of worries | 43 (9.1%) | 30 (7.2%) | 16 (7.4%) | 11 (6.0%) | 15 (13.5%) | 115 (8.2%) | 0.304 |
| Some worries | 129 (27.4%) | 114 (27.2%) | 70 (32.3%) | 47 (25.8%) | 30 (27.0%) | 390 (27.9%) |  |
| No worries | 298 (63.4%) | 275 (65.6%) | 131 (60.4%) | 124 (68.1%) | 66 (59.5%) | 894 (63.9%) |  |
| Total | 470 (100.0%) | 419 (100.0%) | 217 (100.0%) | 182 (100.0%) | 111 (100.0%) | 1399 (100.0%) |  |
| acquaintances from<br>country of origin | 6.4 (7.8) | 7.4 (13.6) | 7.2 (8.6) | 8.4 (11.9) | 6.1 (11.3) | 7.0 (10.8) | 0.142 |
| acquaintances from<br>germany | 6.5 (12.7) | 5.8 (7.8) | 7.1 (11.8) | 5.6 (10.1) | 5.6 (11.3) | 6.2 (10.9) | 0.639 |
| acquaintances from<br>other countries | 4.2 (7.9) | 3.8 (7.2) | 4.4 (8.3) | 4.5 (7.5) | 3.1 (9.6) | 4.0 (7.8) | 0.001 |
| **Sample** |  |  |  |  |  |  |  |
| M3 2016 Flucht (2013-2015) | 124 (26.4%) | 85 (20.2%) | 56 (25.8%) | 32 (17.6%) | 21 (18.9%) | 318 (22.7%) | <0.001 |
| M4 2016 Flucht/Familie (2013-2015) | 142 (30.2%) | 167 (39.8%) | 89 (41.0%) | 46 (25.3%) | 69 (62.2%) | 513 (36.6%) |  |
| M5 2017 Flucht (2013-2016) | 204 (43.4%) | 168 (40.0%) | 72 (33.2%) | 104 (57.1%) | 21 (18.9%) | 569 (40.6%) |  |
| *Total* | *470 (100.0%)* | *420 (100.0%)* | *217 (100.0%)* | *182 (100.0%)* | *111 (100.0%)* | *1400 (100.0%)* |  |
| **Follow-up period** |  |  |  |  |  |  |  |
| 1 year | 239 (50.9%) | 193 (46.0%) | 105 (48.4%) | 124 (68.1%) | 39 (35.1%) | 700 (50.0%) | <0.001 |
| 2 years | 231 (49.1%) | 227 (54.0%) | 112 (51.6%) | 58 (31.9%) | 72 (64.9%) | 700 (50.0%) |  |
| *Total* | *470 (100.0%)* | *420 (100.0%)* | *217 (100.0%)* | *182 (100.0%)* | *111 (100.0%)* | *1400 (100.0%)* |  |

**Table S5:**
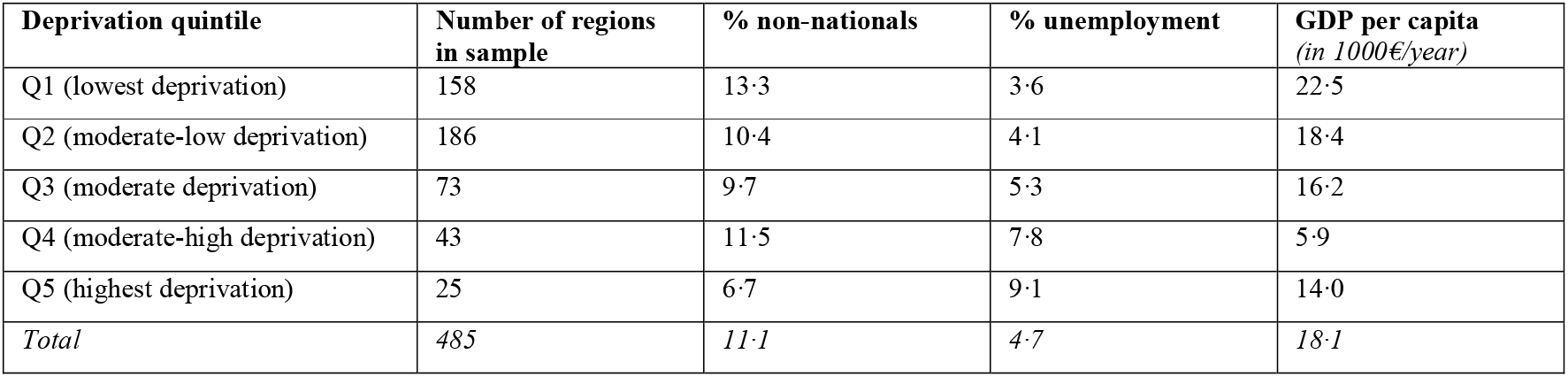
Characteristics of regions covered in the sample.

**Table S6:** Change in mcs and pcs scores between baseline and follow-up by deprivation quintile.

| **Deprivation quintile** | **mcs t0**<br>**mean** | **mcs t1**<br>**mean** | **mcs change**<br>**mean (sd)** | **pcs t0**<br>**mean** | **pcs t1**<br>**mean** | **pcs change**<br>**mean (sd)** | **n** |
| --- | --- | --- | --- | --- | --- | --- | --- |
| Q1 (lowest deprivation) | 48·6 | 47·8 | -0·8 (15·1) | 54·0 | 53·3 | -0·7 (10·3) | 470 |
| Q2 (moderate-low deprivation) | 49·6 | 48·8 | -0·9 (13·8) | 52·6 | 52·8 | 0·3 (10·6) | 420 |
| Q3 (moderate deprivation) | 47·3 | 49·0 | 1·8 (13·1) | 53·3 | 51·9 | -1·4 (9·4) | 217 |
| Q4 (moderate-high deprivation) | 48·9 | 51·4 | 2·4 (14·9) | 52·3 | 50·4 | -1·9 (11·4) | 182 |
| Q5 (highest deprivation) | 47·2 | 50·3 | 3·1 (13·1) | 52·3 | 51·9 | -0·3 (10·1) | 111 |
| *Total* | *48·7* | *49·0* | *0·3 (14·3)* | *53·1* | *52·5* | *-0·6 (10·4)* | *1,400* |

Mcs= mental health component summary scale; pcs= physical health component summary scale

**Table S7:**
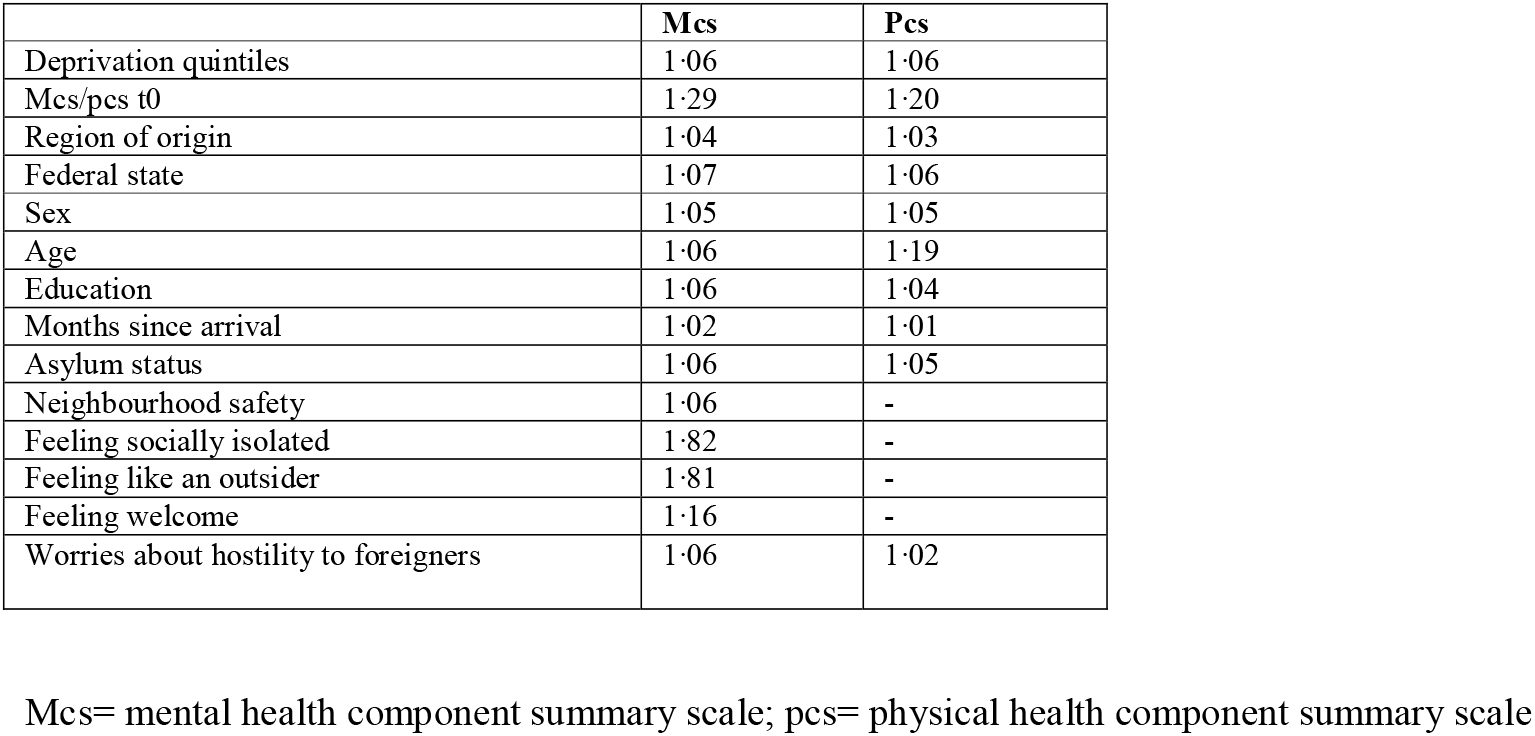
Variance inflation factors of variables included in the final models.

**Table S8:** Association between social context variables and mental/physical health outcomes.

|  | **mcs**<br>mean (sd) | **pcs**<br>mean (sd) | **N** |
| --- | --- | --- | --- |
| **Perception of neighbourhood safety** |  |  |  |
| Very safe | 0.4 (14.3) | -0.5 (10.4) | 1,129.0 |
| Fairly safe | -0.2 (13.5) | -1.0 (10.3) | 221.0 |
| Fairly/ very unsafe | 1.5 (16.7) | -1.6 (11.6) | 50.0 |
| *Total* | *0.3 (14.3)* | *-0.6 (10.4)* | *1,400.0* |
| ANOVA p-value | 0.717 | 0.671 |  |
| **Feeling socially isolated** |  |  |  |
| Very often | 5.4 (18.0) | -0.4 (12.0) | 107.0 |
| Often | 4.6 (15.2) | -1.0 (10.7) | 174.0 |
| Sometimes | -0.5 (13.8) | -0.6 (10.9) | 315.0 |
| Seldom | 0.2 (12.9) | -1.7 (9.4) | 243.0 |
| Never | -1.5 (13.6) | -0.0 (10.1) | 540.0 |
| *Total* | *0.3 (14.3)* | *-0.6 (10.4)* | *1,379.0* |
| ANOVA p-value | <0.001 | 0.298 |  |
| **Feeling like an outsider** |  |  |  |
| Very often | 5.6 (16.9) | 0.1 (11.0) | 118.0 |
| Often | 3.5 (15.4) | -1.1 (10.3) | 181.0 |
| Sometimes | 0.7 (14.1) | -0.8 (10.6) | 373.0 |
| Seldom | 0.0 (13.3) | -1.2 (10.3) | 267.0 |
| Never | -2.7 (13.3) | -0.1 (10.2) | 426.0 |
| *Total* | *0.3 (14.4)* | *-0.6 (10.4)* | *1,365.0* |
| ANOVA p-value | <0.001 | 0.541 |  |
| **Feeling welcome** |  |  |  |
| Completely | -0.4 (14.4) | -0.4 (10.4) | 885.0 |
| For the most part | 0.7 (13.8) | -1.0 (10.7) | 272.0 |
| In some respects | 2.4 (14.0) | -1.4 (10.4) | 164.0 |
| Hardly/ not at all | 3.5 (14.8) | -0.3 (9.8) | 73.0 |
| *Total* | *0.3 (14.3)* | *-0.6 (10.4)* | *1,394.0* |
| ANOVA p-value | 0.020 | 0.576 |  |
| **Worries about hostility towards foreigners** |  |  |  |
| A lot of worries | -3.2 (15.1) | -1.3 (10.9) | 115.0 |
| Some worries | -1.5 (13.8) | -0.3 (10.2) | 390.0 |
| No worries | 1.6 (14.2) | -0.7 (10.4) | 894.0 |
| *Total* | *0.3 (14.3)* | *-0.6 (10.4)* | *1,399.0* |
| ANOVA p-value | <0.001 | 0.639 |  |
| **No. of acquaintances from country of origin (tertiles)** |  |  |  |
| 1 (lowest no. of acquaintances) | 0.6 (14.0) | -0.5 (10.5) | 522.0 |
| 2 | -0.8 (15.1) | 0.1 (9.7) | 335.0 |
| 3 (highest no. of acquaintances) | 1.7 (13.7) | -1.4 (10.9) | 381.0 |
| *Total* | *0.5 (14.2)* | *-0.6 (10.4)* | *1,238.0* |
| ANOVA p-value | 0.052 | 0.141 |  |
| **No. of acquaintances from Germany (tertiles)** |  |  |  |
| 1 (lowest no. of acquaintances) | 0.5 (14.4) | -0.7 (10.7) | 475.0 |
| 2 | -0.9 (15.0) | -0.5 (9.4) | 282.0 |
| 3 (highest no. of acquaintances) | 1.0 (13.6) | -0.8 (10.3) | 339.0 |
| *Total* | *0.3 (14.3)* | *-0.7 (10.3)* | *1,096.0* |
| ANOVA p-value | 0.215 | 0.911 |  |
| **No. of acquaintances from other countries (tertiles)** |  |  |  |
| 1 (lowest no. of acquaintances) | 0·9 (14·3) | -0·3 (10·7) | 372·0 |
| 2 | -0·6 (15·3) | -0·4 (9·9) | 301·0 |
| 3 (highest no. of acquaintances) | 1·1 (13·2) | -1·1 (10·0) | 310·0 |
| *Total* | *0·5 (14·3)* | *-0·6 (10·2)* | *983·0* |
| ANOVA p-value | 0·237 | 0·556 |  |
| **Type of accommodation** |  |  |  |
| Shared accommodation | 0·6 (15·3) | -0·4 (10·3) | 556 |
| Private accommodation | 0·1 (13·6) | -0·9 (10·5) | 841 |
| *Total* | *0·3 (14·3)* | *-0·7 (10·4)* | *1397* |
| ANOVA p-value | 0·554 | 0·399 |  |

Mcs= mental health component summary scale; pcs= physical health component summary scale

**Table S9:** Outcomes of mediation analyses mcs/pcs.

|  | Mcs (t1-t0) |  | Pcs (t1-t0) |  |
| --- | --- | --- | --- | --- |
|  | **Sociodemo + social**<br>**mediation variables** | **Sociodemo +**<br>**accommodation** | **Sociodemo + social**<br>**mediation variables** | **Sociodemo +**<br>**accommodation** |
| clustering | commune | commune | none | none |
| Q1<br>(lowest deprivation) | ref | ref | ref | ref |
| Q2<br>(moderate-low<br>deprivation) | -0.12299<br>(-1.70699 - 1.46100) | 0.06382<br>(-1.55041 - 1.67805) | -0.20372<br>(-1.44901 - 1.04157) | -0.11506<br>(-1.33377 - 1.10364) |
| Q3<br>(moderate deprivation) | 0.42185<br>(-1.65646 - 2.50017) | -0.02223<br>(-2.25138 - 2.20692) | -0.88677<br>(-2.56348 - 0.78994) | -0.85963<br>(-2.40965 - 0.69038) |
| Q4<br>(moderate-high<br>deprivation) | 2.19674<br>(-0.62327 - 5.01676) | 2.22937<br>(-0.92549 - 5.38422) | -2.37190*<br>(-4.28690 - -0.45690) | -2.10171*<br>(-4.03228 - -0.17115) |
| Q5<br>(high deprivation) | 1.55826<br>(-3.70747 - 6.82400) | 1.96715<br>(-3.45347 - 7.38778) | -0.84332<br>(-4.19508 - 2.50845) | -0.94794<br>(-4.02151 - 2.12563) |
| Very safe neighbourhood | ref |  | ref |  |
| Fairly safe neighbourhood | -2.04808*<br>(-4.04376 - -0.05241) |  | -0.51231<br>(-1.92841 - 0.90379) |  |
| Fairly/ very unsafe<br>neighbourhood | -0.87344<br>(-4.75285 - 3.00596) |  | -1.55240<br>(-4.40718 - 1.30238) |  |
| Very often socially<br>isolated | ref |  | ref |  |
| Often socially isolated | -3.61703+<br>(-7.39456 - 0.16050) |  | 0.65786<br>(-1.94368 - 3.25941) |  |
| Sometimes socially<br>isolated | -2.25032+<br>(-4.90468 - 0.40403) |  | -0.29951<br>(-2.18206 - 1.58305) |  |
| Seldom socially isolated | -3.61255**<br>(-5.57926 - -1.64584) |  | -0.59972<br>(-2.23623 - 1.03678) |  |
| Never socially isolated | -1.19715<br>(-3.02192 - 0.62762) |  | -0.50793<br>(-2.13355 - 1.11768) |  |
| Very often feeling like an<br>outsider | ref |  | ref |  |
| Often feeling like an<br>outsider | -0.83323<br>(-4.17505 - 2.50860) |  | -1.67519<br>(-3.94440 - 0.59402) |  |
| Sometimes feeling like an<br>outsider | -0.84832<br>(-3.36475 - 1.66812) |  | -0.68009<br>(-2.48260 - 1.12243) |  |
| Seldom feeling like an<br>outsider | 0.90524<br>(-0.97446 - 2.78494) |  | -0.78199<br>(-2.36502 - 0.80104) |  |
| Never feeling like an<br>outsider | 1.31136<br>(-0.50037 - 3.12310) |  | -0.20139<br>(-1.80650 - 1.40373) |  |
| A lot of worries about<br>hostility to foreigners | ref |  | ref |  |
| Some worries about<br>hostility to foreigners | -6.05757**<br>(-8.79279 - -3.32234) |  | -2.13110*<br>(-4.05194 - -0.21026) |  |
| No worries about hostility<br>to foreigners | -4.12526**<br>(-5.52350 - -2.72702) |  | -0.36027<br>(-1.55295 - 0.83240) |  |
| Shared accommodation |  | ref |  | ref |
| Private accommodation |  | 1.10424<br>(-0.46138 - 2.66987) |  | -0.09924<br>(-1.17572 - 0.97723) |
| Observations | 1191 | 1225 | 1191 | 1225 |
| R-squared |  |  | 0.3743239 | 0.3654582 |
| adjusted R-squared |  |  | 0.3508679 | 0.3484235 |
| model degrees of freedom | 43 | 32 | 43 | 32 |
| Number of clusters | 431 | 439 |  |  |
| Aikikes information criterion (AIC) | 9051.178 | 9360.816 |  |  |
| Bayes information criterion (BIC) | 9284.975 | 9539.69 |  |  |
| Intra-class correlation coefficient (ICC) | 0.052187 | 0.0822581 |  |  |
| Neighbourhood intercept variance | 5.685784 | 9.61043 |  |  |
| Residual variance | 103.2645 | 107.2223 |  |  |
| Proportional change in variance vs. null model (PCV<sub>N</sub>) |  |  |  |  |
| Likelihood-ratio test vs. linear model | 0.006 | 0.0003 |  |  |

All models adjusted for baseline and socio-demographic variables: baseline mcs/pcs, federal state, region of origin, sex, age, education, asylum status and months since arrival in Germany
Mcs= mental health component summary scale; pcs= physical health component summary scale
Significance levels: + $p < 0.1$ ; \* $p < 0.05$ ; \*\* $p < 0.01$

**Table S10:** Outcomes of sensitivity analyses mcs.

|  | **Mcs (t1-t0)** |  |  |  |  |  |
| --- | --- | --- | --- | --- | --- | --- |
|  | **s1** | **s2** | **s3** | **s4** | **s5** | **s6** |
| Clustering | commune | commune | commune | commune | commune | commune |
| Q1<br>(lowest deprivation) | ref | ref | ref | ref | ref | ref |
| Q2<br>(moderate-low deprivation) | -0.36987<br>(-1.91238 - 1.17265) | -0.15799<br>(-1.76120 - 1.44521) | -0.05435<br>(-1.73999 - 1.63130) | -0.35855<br>(-1.94979 - 1.23269) | -0.12225<br>(-1.72578 - 1.48129) | -.6533268<br>(-2.497691 - 1.191037) |
| Q3<br>(moderate deprivation) | 0.69497<br>(-1.24903 - 2.63897) | 0.36492<br>(-1.79405 - 2.52390) | 0.07508<br>(-2.12202 - 2.27218) | 0.21556<br>(-1.74357 - 2.17470) | 0.40246<br>(-1.76615 - 2.57108) | .1774576<br>(-2.175607 - 2.530522) |
| Q4<br>(moderate-high deprivation) | 2.16982+<br>(-0.33313 - 4.67276) | 2.24817<br>(-0.58596 - 5.08230) | 2.11455<br>(-0.99616 - 5.22526) | 1.76467<br>(-0.62081 - 4.15015) | 2.20018<br>(-0.63716 - 5.03752) | .4320928<br>(-2.256505 - 3.120691) |
| Q5<br>(high deprivation) | 3.11849<br>(-1.25989 - 7.49687) | 1.50883<br>(-3.67844 - 6.69611) | 1.30828<br>(-3.97720 - 6.59377) | 1.10446<br>(-4.28634 - 6.49526) | 1.54688<br>(-3.64591 - 6.73967) | -.3523031<br>(-4.12403 - 3.419424) |
| Years of follow-up |  | 0.55862<br>(-0.88652 - 2.00376) |  |  |  |  |
| Survey wave |  |  |  |  | -0.08357<br>(-1.02274 - 0.85559) |  |
| Observations | 1146 | 1191 | 1133 | 1170 | 1191 | 1065 |
| Number of groups | 417 | 431 | 416 | 430 | 431 | 425 |

All models adjusted for baseline and socio-demographic variables: baseline mcs, federal state, region of origin, sex, age, education, asylum status and months since arrival in Germany as well as social network variables (neighbourhood safety, feeling socially isolated, feeling like an outsider, worries about hostility to foreigners)
Mcs= mental health component summary scale
Significance levels: + p<0.1; \*p<0.05; \*\*p<0.01

**Table S11:** Outcomes of sensitivity analyses pcs.

|  | Pcs (t1-t0) |  |  |  |  |  |
| --- | --- | --- | --- | --- | --- | --- |
|  | s1 | s2 | s3 | s4 | s5 | s6 |
| Clustering | none | none | none | none | none | none |
| Q1<br>(lowest deprivation) | ref | ref | ref | ref | ref | ref |
| Q2<br>(moderate-low deprivation) | 0.09964<br>(-1.14154 - 1.34083) | -0.11398<br>(-1.33970 - 1.11174) | -0.00723<br>(-1.24738 - 1.23293) | -0.20086<br>(-1.34344 - 0.94171) | -0.10560<br>(-1.32818 - 1.11697) | -0.5561305<br>(-1.851517 - 0.739256) |
| Q3<br>(moderate deprivation) | -1.06741<br>(-2.70311 - 0.56830) | -0.91689<br>(-2.45476 - 0.62098) | -0.99841<br>(-2.74142 - 0.74459) | -1.12269<br>(-2.66586 - 0.42048) | -0.81954<br>(-2.36387 - 0.72480) | -2.013988 *<br>(-3.838724 - -0.1892521) |
| Q4<br>(moderate-high deprivation) | -2.03772*<br>(-3.95316 - -0.12228) | -2.14412*<br>(-4.09354 - -0.19471) | -2.37481*<br>(-4.38881 - -0.36080) | -1.27125<br>(-2.93830 - 0.39580) | -2.15732*<br>(-4.10468 - -0.20997) | -2.930691 **<br>(-5.086605 - -0.7747778) |
| Q5<br>(high deprivation) | -1.17374 | -0.99068<br>(-1.03119 - 1.22347) | -0.55116 | -0.83058 | -0.93190 | -1.77103<br>(-4.934023, 1.391962) |
| Years of follow-up |  | 0.09614<br>(-1.03119 - 1.22347) |  |  |  |  |
| Survey wave |  |  |  |  | 0.26959<br>(-0.42821 - 0.96739) |  |
| Observations | 1180 | 1226 | 1165 | 1200 | 1226 | 1099 |
| R-squared | 0.35441 | 0.36558 | 0.37381 | 0.32407 | 0.36586 | 0.3532 |

All models adjusted for baseline and socio-demographic variables: baseline pcs, federal state, region of origin, sex, age, education, asylum status and months since arrival in Germany
pcs= physical health component summary scale
Significance levels: + $p < 0.1$ ; \* $p < 0.05$ ; \*\* $p < 0.01$

**Figure S12:**
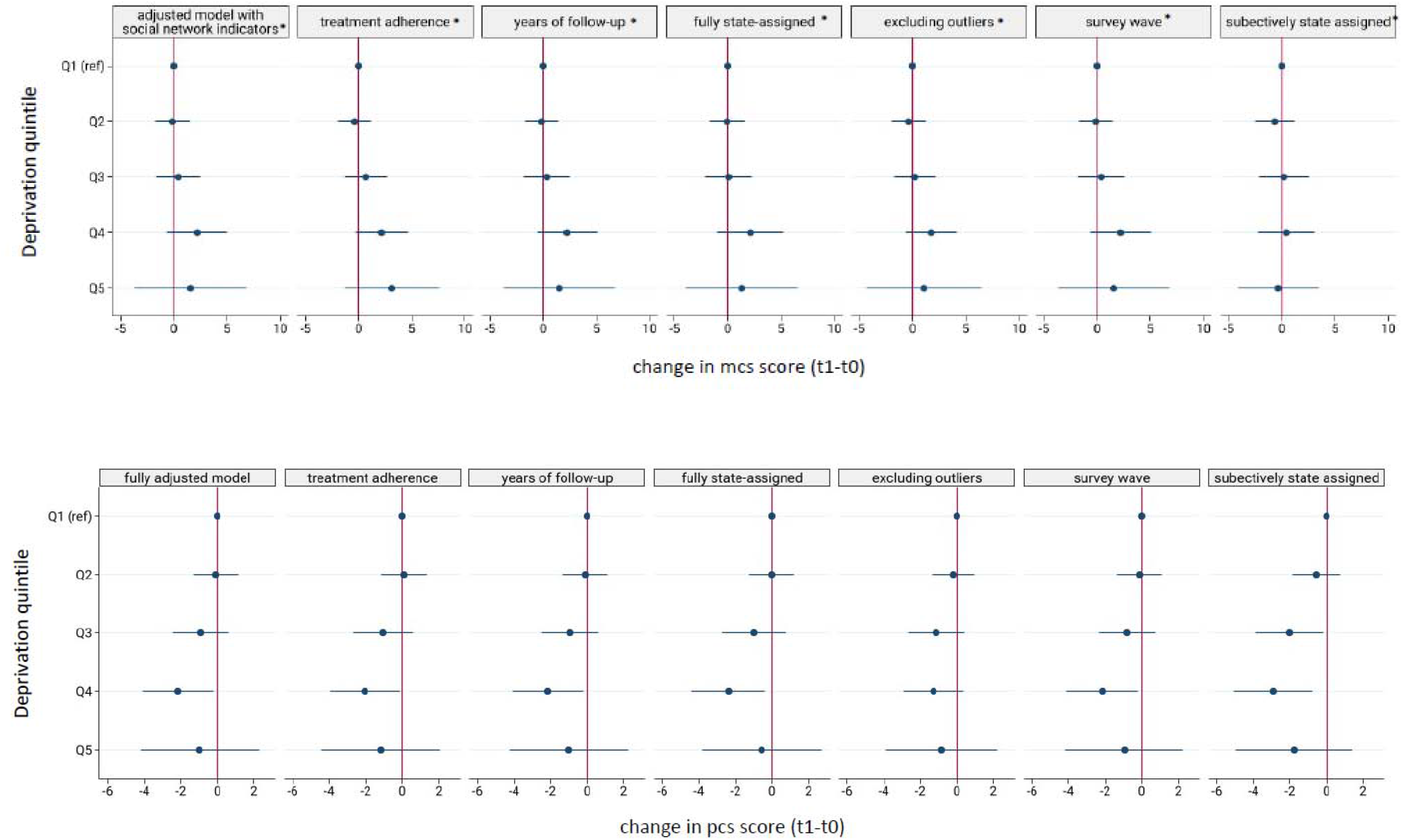
Outcomes of sensitivity analyses mcs/pcs.

**Table S13:**
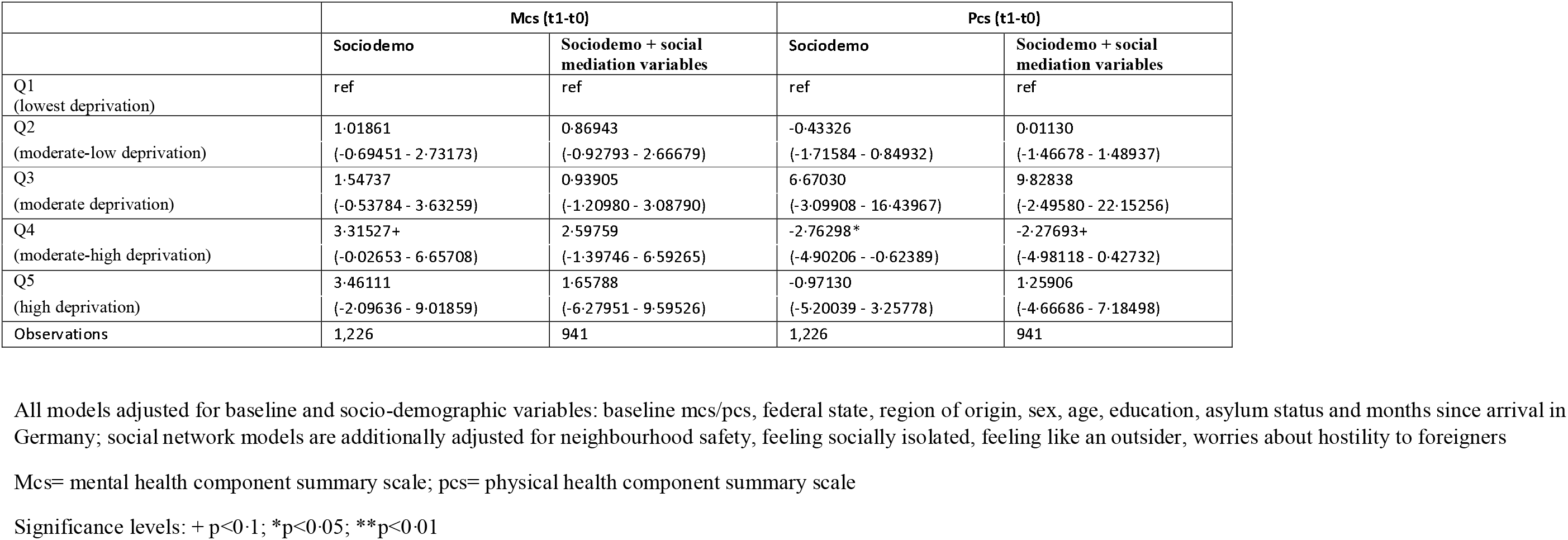
Outcomes of sensitivity analysis s7: average treatments effects (ATE) estimated with inverse proportional weighting for mcs and pcs.

|  | Mcs (t1-t0) |  | Pcs (t1-t0) |  |
| --- | --- | --- | --- | --- |
|  | Sociodemo | Sociodemo + social mediation variables | Sociodemo | Sociodemo + social mediation variables |
| Q1<br>(lowest deprivation) | ref | ref | ref | ref |
| Q2<br>(moderate-low deprivation) | 1.01861<br>(-0.69451 - 2.73173) | 0.86943<br>(-0.92793 - 2.66679) | -0.43326<br>(-1.71584 - 0.84932) | 0.01130<br>(-1.46678 - 1.48937) |
| Q3<br>(moderate deprivation) | 1.54737<br>(-0.53784 - 3.63259) | 0.93905<br>(-1.20980 - 3.08790) | 6.67030<br>(-3.09908 - 16.43967) | 9.82838<br>(-2.49580 - 22.15256) |
| Q4<br>(moderate-high deprivation) | 3.31527+<br>(-0.02653 - 6.65708) | 2.59759<br>(-1.39746 - 6.59265) | -2.76298*<br>(-4.90206 - -0.62389) | -2.27693+<br>(-4.98118 - 0.42732) |
| Q5<br>(high deprivation) | 3.46111<br>(-2.09636 - 9.01859) | 1.65788<br>(-6.27951 - 9.59526) | -0.97130<br>(-5.20039 - 3.25778) | 1.25906<br>(-4.66686 - 7.18498) |
| Observations | 1,226 | 941 | 1,226 | 941 |

All models adjusted for baseline and socio-demographic variables: baseline mcs/pcs, federal state, region of origin, sex, age, education, asylum status and months since arrival in Germany; social network models are additionally adjusted for neighbourhood safety, feeling socially isolated, feeling like an outsider, worries about hostility to foreigners
Mcs= mental health component summary scale; pcs= physical health component summary scale
Significance levels: + p<0.1; \*p<0.05; \*\*p<0.01

**Table S14:**
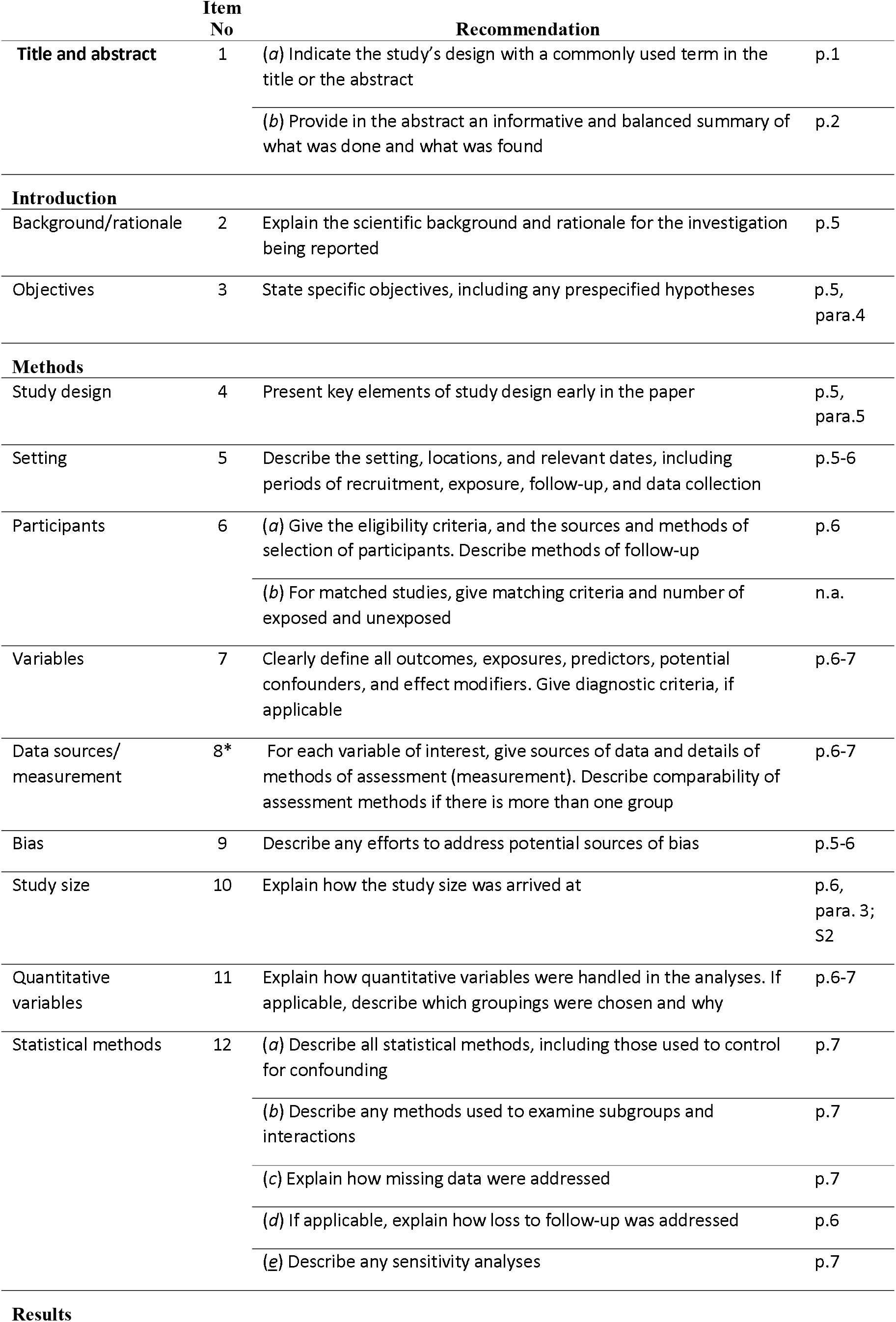
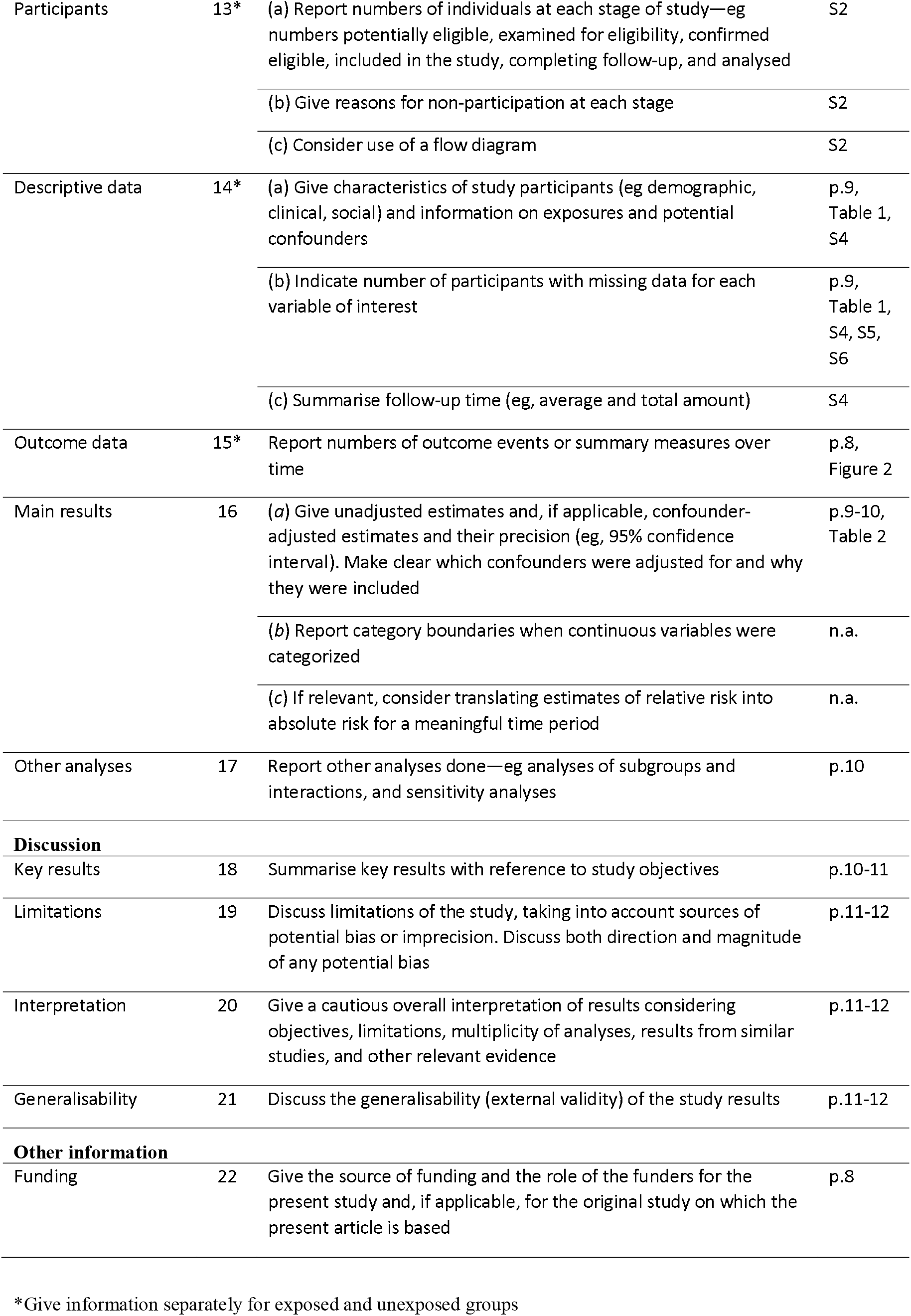
STROBE Statement for cohort studies.

